# COVID-19 optimal vaccination policies: a modeling study on efficacy, natural and vaccine-induced immunity responses

**DOI:** 10.1101/2020.11.19.20235176

**Authors:** Manuel Adrian Acuña-Zegarra, Saúl Díaz-Infante, David Baca-Carrasco, Daniel Olmos Liceaga

## Abstract

At the date, Europe and part of North America face the second wave of COVID-19, causing more than 1 300 000 deaths worldwide. Humanity lacks successful treatments, and a sustainable solution is an effective vaccine. Pfizer and the Russian Gamaleya Institute report that its vaccines reach more than 90 % efficacy in a recent press release. If third stage trial results favorable, pharmaceutical firms estimate big scale production of its vaccine candidates around the first 2021 quarter and the World Health organization fix as objective, vaccinate 20 % of the whole population at the final of 2021. However, since COVID-19 is new to our knowledge, vaccine efficacy and induced-immunity responses remain poorly understood. There are great expectations, but few think the first vaccines will be fully protective. Instead, they may reduce the severity of illness, reducing hospitalization and death cases.

Further, logistic supply, economic and political implications impose a set of grand challenges to develop vaccination policies. For this reason, health decision-makers require tools to evaluate hypothetical scenarios and evaluate admissible responses.

Our contribution answers questions in this direction. According to the WHO Strategic Advisory Group of Experts on Immunization Working Group on COVID-19 Vaccines, we formulate an optimal controlled model to describe vaccination policies that minimize the burden of COVID-19 quantified by the number of disability-adjusted years of life lost. Additionally, we analyze the reproductive vaccination number according to vaccination profiles depending on coverage, efficacy, horizon time, and vaccination rate. We explore scenarios regarding efficacy, coverage, vaccine-induced immunity, and natural immunity via numerical simulation. Our results suggest that response regarding vaccine-induced immunity and natural immunity would play a dominant role in the vaccination policy design. Likewise, the vaccine efficacy would influence the time of intensifying the number of doses in the vaccination policy.

## 1. Introduction

In late December 2019, a new virus’s appearance is reported in Wuhan City, Hubei Province, China. Called SARS-CoV2, it is the virus that causes the 2019 coronavirus disease (COVID-19) and that, very quickly since its appearance, has spread throughout much of the world, causing severe problems to health systems of all the countries in which it is present [1].

Due to the absence of successful treatment and vaccines, several non-pharmaceutical interventions (NPIs) have been implemented in all the countries where the disease is present, with quarantine, isolation, and social distancing being the main ones [2, 3]. Despite the measures that different governments have taken to mitigate the epidemic, it has not been controlled in most places, which may be due to the relaxation of mitigation measures. At the date of writing this work, the upturn or regrowth in the number of cases in some countries around the world has been observed. In some places, this behavior is referred to as “second wave”. On the other hand, since the new coronavirus appearance, the international scientific community has been working to understand the virus nature. They mainly focus on the spreading mechanisms between individuals, and developing vaccines and treatments to reduce the number of infections and fatality cases.

Development of COVID-19 vaccines is the major challenge of these days. Some research efforts in this direction can be found in [4, 5]. In Mexico, some of the considered vaccines to be applied to the population are Adenovirus Type 5 Vector (Ad5-nCoV) by Cansino Biologics, AZD1222 by AstraZeneca and BNT162b2 by Pfizer and BioNTech. At the current date, these and other vaccines are on the testing phase (phase 3) and it is believed that their distribution will begin at the end of March 2021. However, there are still somequestions about the efficacy of the vaccines. The U. S. Food and Drugs Administration (FDA) requires firm evidence that a vaccine protects at least half of those inoculated [6]. At the date, there are no conclusive results about the vaccines’ efficacy, nor the immunity time induced by vaccines.

To get a clearer understanding of different vaccination strategies and their consequences on the number of infected individuals, Kermack–McKendrick type models have taken a leading role. This kind of models have been used to understand vaccination dynamics on other diseases [7]. It is important to stress that nowadays Kermack–McKendrick type mathematical models has helped to describe COVID-19 epidemics properties around the globe. These models have been used to estimate the basic reproductive number associated with the disease and also different parameters involved in its spread [8, 9]. Another use of this kind of model has been addressed to propose and evaluate the effect of various control measures classified as NPIs [8, 10–14].

On the other hand, vaccines’ development for COVID-19 has led to ask which are the best vaccination strategies to reduce the disease levels in a population. In order to find a good vaccination strategy, a widely used tool is the optimalcontrol theory. This mathematical method together with Kermack–McKendrick models are useful to propose scenarios in which the vaccine’s application minimizes the damage caused by the disease and its application cost. Some results about it have been applied to control other diseases for humans and animals [15–18]. Optimal control theory has also been used in COVID-19 studies. Most efforts have been invested in finding optimal strategies to evaluate the impact of non-pharmaceutical interventions [19–21]. Optimal control strategies have also being considered in vaccination strategies [22]. In this work, the authors took their optimization based on an extension of the SIR mathematical model for COVID-19, which included a vaccinated class. Within their objectives was to minimize only the number of infected individuals together with the prescribed vaccine concentration during treatment.

In this work, we present a mathematical model to describe the transmission and some vaccination dynamics of COVID-19. The aim is to implement optimal control theory to obtain vaccination strategies in a homogeneous population. Such strategies are aligned to the policies of the WHO strategic advisory group of experts (SAGE) on COVID-19 vaccination [23]. In this sense we look to minimize the disability-adjusted life year (DALY) [24]. This quantity is used by WHO to quantify the burden of disease from mortality and morbidity, which is given by the sum of the years of life lost (YLL) and years lost due to disability (YLD). Motivated by this, we arrive to a different optimal control problem as the one studied in [22]. In particular, we focus on vaccination efficacy, natural immunity and a given coverage for a particular horizon time, to find an optimal daily vaccination rates strategy. By following this idea, we can (i) find strategies such that hospitals are not surpassed in their capacity, (ii) value a strategy by obtaining estimates of the number of infected and deceased individuals and (iii) make comparisons between a constant vaccination policy versus the optimal vaccination policy.

Our manuscript is divided into the following sections. In Section 2, we present our mathematical model, which includes preventive vaccine dynamics. Section 3 includes an analysis of the vaccination reproductive number. In Section 4, we define our optimal control problem when modulating the vaccination rate by a time-dependent control signal *u*_*V*_ (*t*). Section 5 presents our numerical results regarding optimal vaccination policies. We end with a conclusions and discussion section.

## 2. Mathematical model formulation

In this section, we formulate our baseline mathematical model, which additionally to the transmission dynamics, includes vaccination. In order to build our model, we follow the classical Kermack-McKendrick approach. Figure 1 shows the compartmental diagram of our mathematical model.

**Figure 1:**
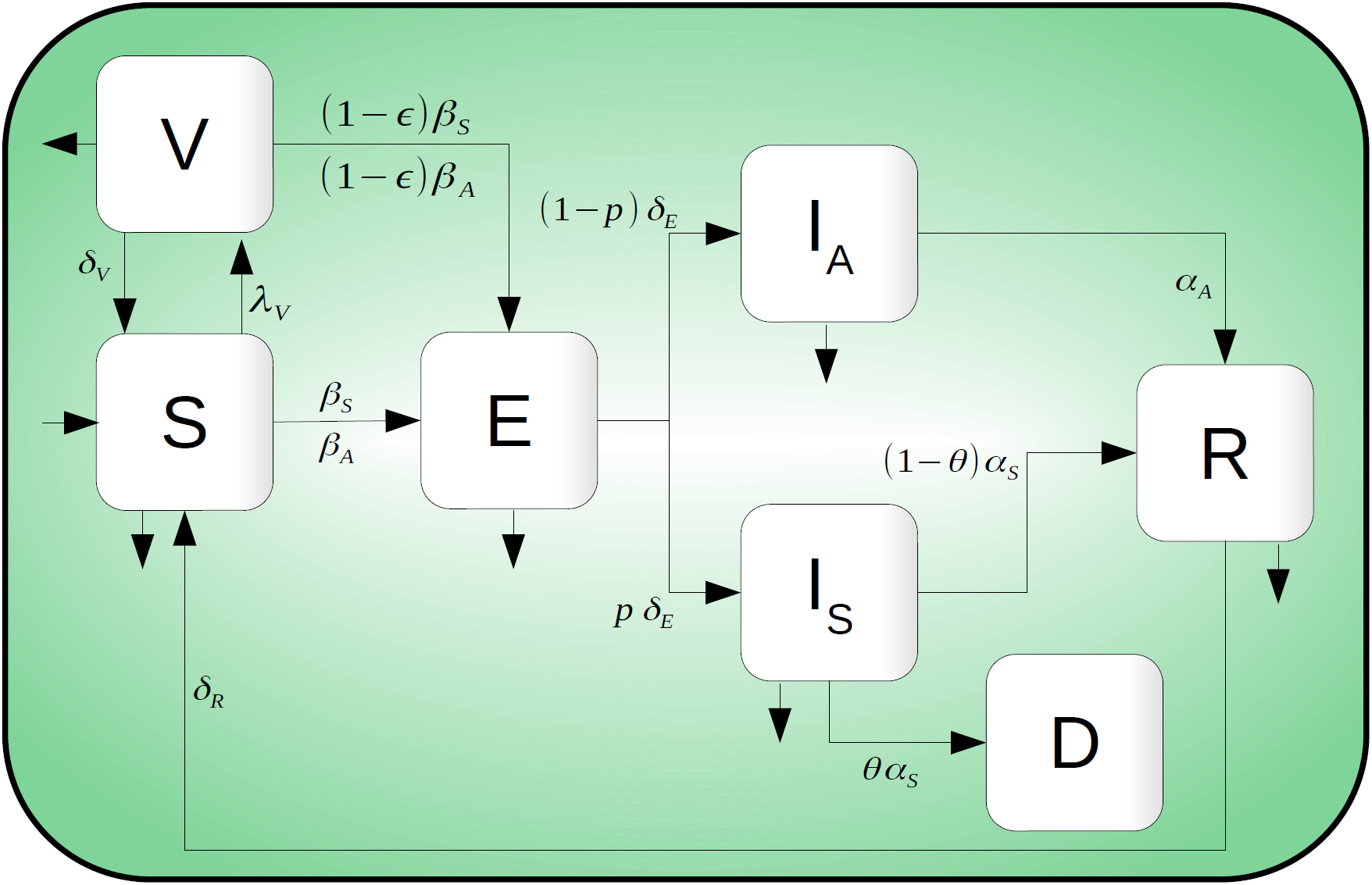
Compartmental diagram of COVID-19 transmission dynamics which including vaccination dynamics. Here, there are seven different classes: Susceptible (*S*), exposed (*ϵ*), symptomatic infected (*I*_*S*_), asymptomatic infected (*I*_*A*_), recovered (*R*), death (*D*) and vaccinated (*V*) individuals. It is important to mention that *I*_*S*_ represents the proportion of symptomatic individuals who will later report to some health medical center.

Information about reinfection dynamics on COVID-19 disease is unclear to date. However, to explore some scenarios related to this dynamic, we assume that reinfection is possible after a period of time. On the other hand, we model the vaccination process considering some assumptions: i) Vaccination is applied to all the alive individuals except those in the symptomatic class. In this situation, vaccines are applied indiscriminately over individuals on the *S, E, I*_*A*_, and *R* classes; ii) the vaccine has preventive nature, that is, only reflected in the susceptible individuals (*S*); iii) people will only get one vaccine during the campaign, and iv) vaccines do not necessarily have a hundred percent of effectivity, which implies that some vaccinated people can get the disease. We denote the effectivity rate by *ϵ*. Based on these assumptions our model becomes

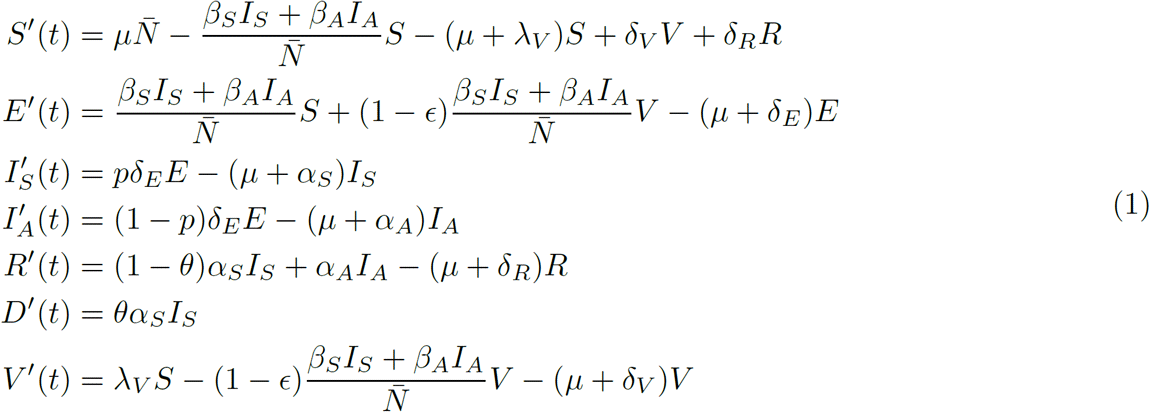

where 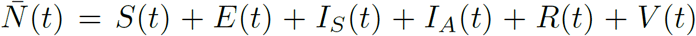 and 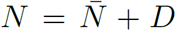. Additionally, we include the equations

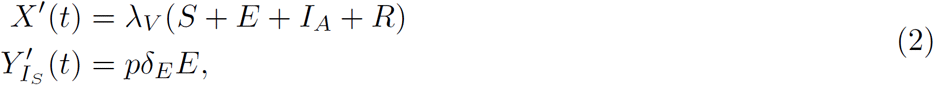

where *X*(*t*) and 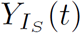 represent the cumulative doses at time *t*, and the cumulative incidence at time *t*, respectively. Parameters description of system in Equation (1) is provided in Table 1.

**Table 1:**
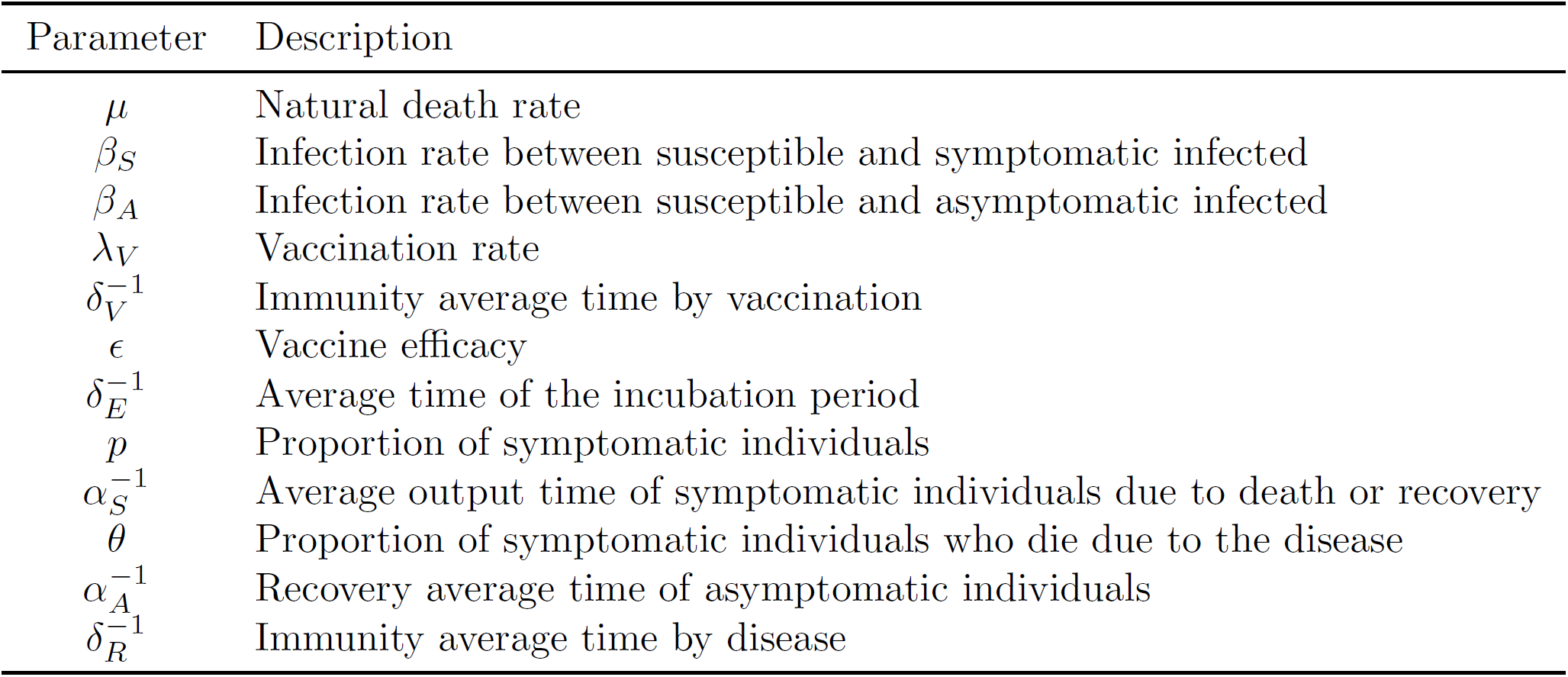
Parameters definition of system in Equation (1).

### 2.1 Baseline parameters value and initial conditions

It is now necessary to define a set of baseline parameter values to explore some scenarios of interest. In the present work, we consider Mexico City plus Mexico state as our study region and use COVID-19 data to estimate some parameter values. We follow a Bayesian approach to address this problem. We use a negative binomial distribution as an observation model, in which the mean parameter is given by

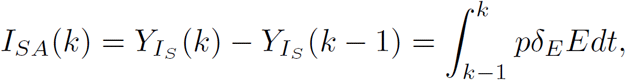

where *I*_*SA*_(*k*) represents the incidence per day of infected symptomatic individuals at the *k − th* day. The parameter estimation splits into two-stage: before and after mitigation measures were implemented, and we only focus on the early phase of the COVID-19 outbreak. Figure 2 shows fitting curves with their respective confidence bands for both stages. Here, we observe that our estimations follow the growth profile of the epidemic curve. For more information about the parameter estimation process, see Appendix A. Table 2 resume our parameter calibration. On the other hand, since it is unclear when the vaccines will be available, then we assume that our scenarios start on the growth stage of a second outbreak (see Figure 3) with the objective of starting under a plausible scenario. Thus, for numerical results, initial conditions normalized with total population *N* = 26 446 435, result *S*(0) = 0.463 606 046 009 872, *E*(0) = 0.000 670 33, *I*_*S*_(0) = 0.000 092 83, *I*_*A*_(0) = 0.001 209 86, *R*(0) = 0.532 520 194, *D*(0) = 0.001 900 74, *V* (0) = 0, *X*(0) = 0, and 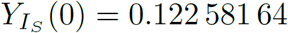.

**Table 2:** Confidence interval for some parameters of system in Equation (1), and for the basic reproductive number (*R*_0_).

| Parameter | 95% Confidence Interval |
| --- | --- |
| $\beta_S$ | [0.248 371 2, 0.487 207 14] |
| $\beta_A$ | [0.185 169 6, 0.321 812 49] |
| $p$ | [0.061, 0.2206] |
| $R_0$ | [1.702, 1.887] |

**Figure 2:**
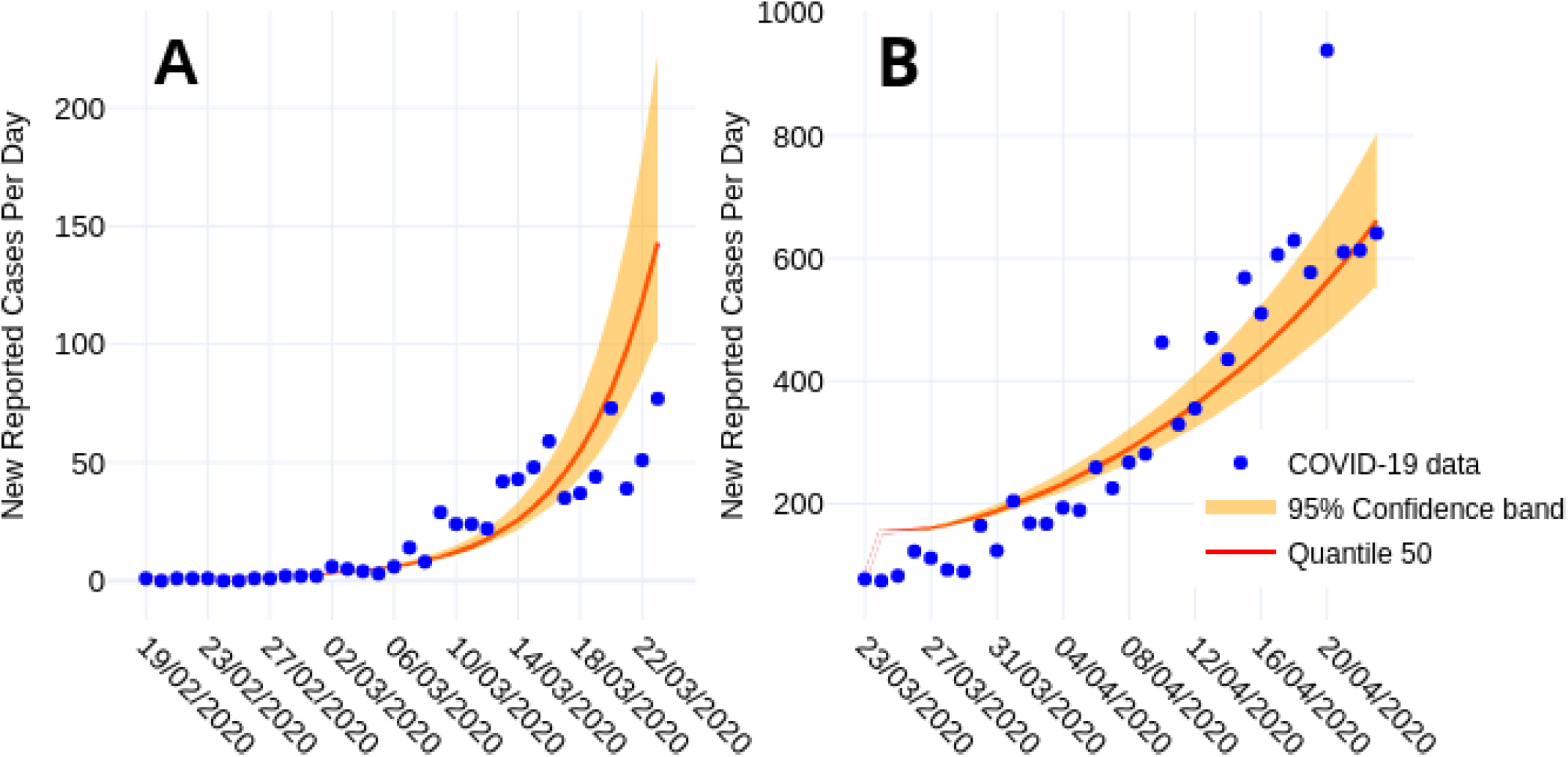
Fitting curves for the early phase of the COVID-19 outbreak in Mexico City plus Mexico state. (A) Outbreak from February 19 to March 23, 2020. (B) Outbreak from March 23 to April 23, 2020. Reported data are shown in blue points, solid red line denote quantile 50 of all *I*_*SA*_ solutions.

**Figure 3:**
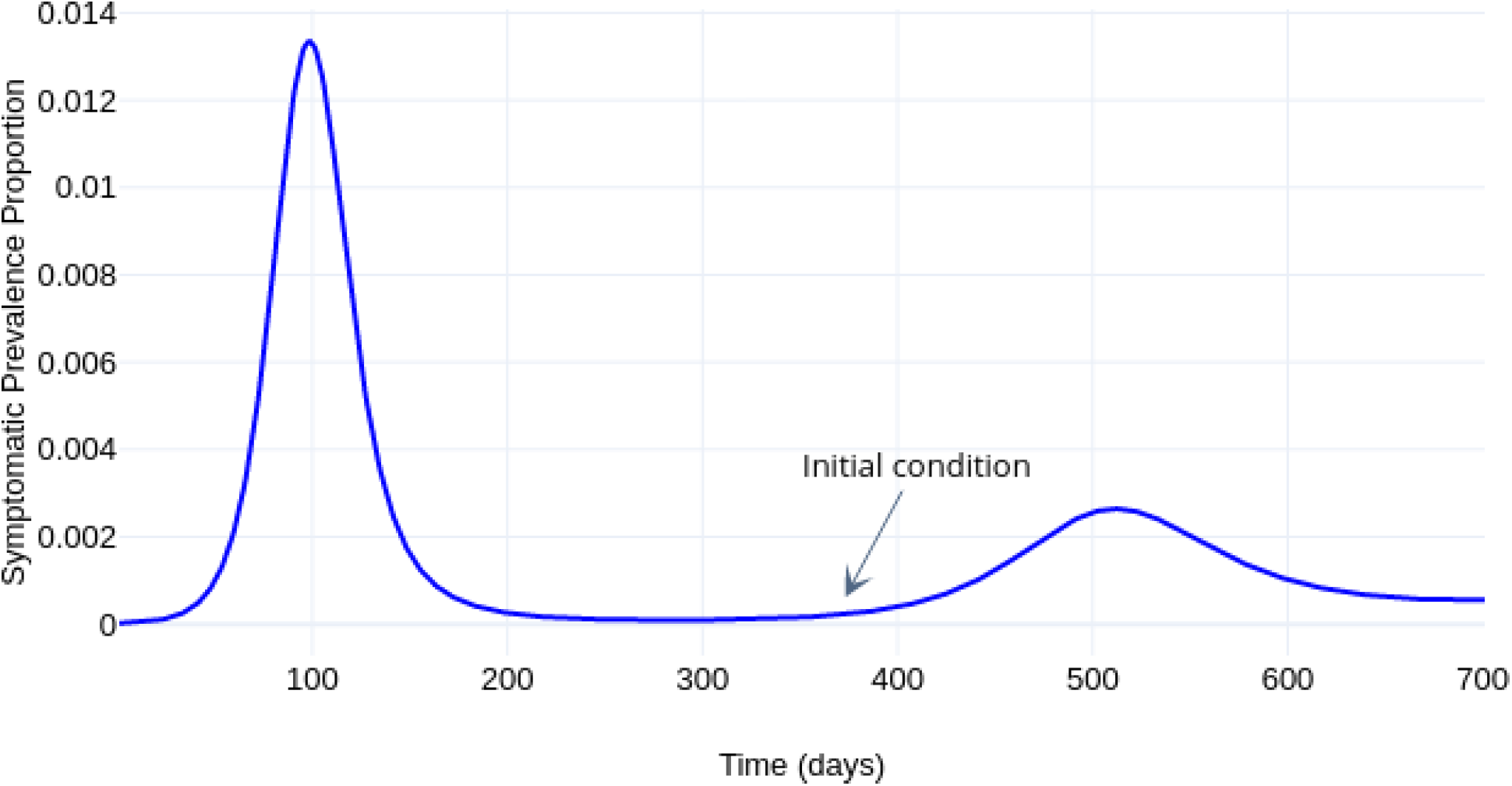
Dynamics of the symptomatic individuals without vaccination. Black arrow indicates the outbreak stage in which we start our control simulations.

## 3. Model Analysis

In this section, we use the estimated parameters from the previous section to analyze model in Equation (1) and the effect of the vaccine. Some plausible scenarios are presented, depending on the effectiveness of the vaccine, as well as the rate of vaccination.

In the first instance, it has been shown that the interest region of the state variables of the system is positively invariant and the proof can be found in Appendix B.

Now, an important concept in the analysis of the spread of diseases is the basic reproductive number, defined as the number of secondary infections produced by a typical infected individual, throughout his infectious period, when in contact with a totally susceptible population. Following ideas of [25], the basic reproduction number for system in Equation (1) is (see Appendix B)

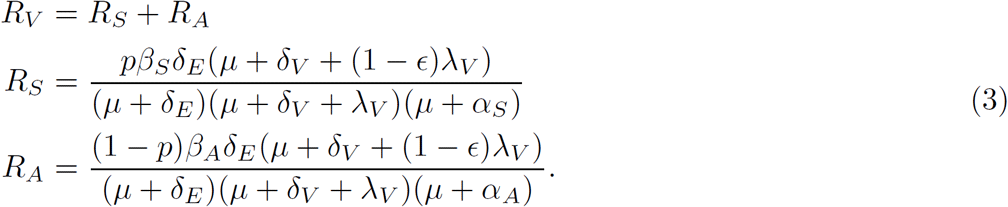

Note that each sum of *R*_*V*_ represents the contribution of the symptomatic and asymptomatic infected, respectively, to the spread of the disease. Following the ideas of Alexander et. al. [7], expression for *R*_*V*_ can be rewritten as

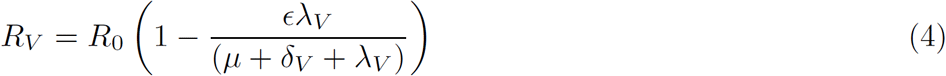

where

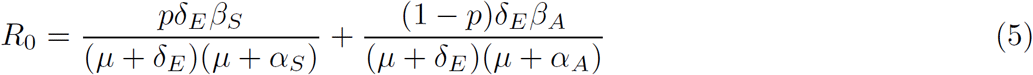

is the basic reproduction number of the system without vaccine. Note that 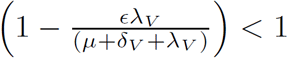. Therefore, this factor, which enclose the parameters corresponding to the application of the vaccine, will allow us to modulate the value of *R*_0_. In the first instance, if *R*_0_ *<* 1, then *R*_*V*_ *<* 1. But, if *R*_0_ *>* 1, we ask if the application of the vaccine can lower *R*_*V*_ value below 1. In this sense, it is easy to prove that, if

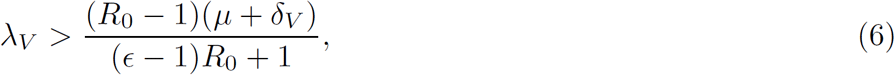

for *ϵ >* 1 *−* (1*/R*_0_), it is possible to reduce the value of *R*_*V*_ below one. That is, there is a region in the parameter space in which it is possible to reduce the value of *R*_*V*_ below one, considering adequate efficacy, vaccination rate and duration of the effect of the vaccine. However, if the Inequality (6) is not satisfied, it will not be possible to reduce the value of *R*_*V*_ below 1.

To illustrate the aforementioned, Figure 4 shows the regions where it is possible to reduce the value of *R*_*V*_. In this case, we set all the system parameters as given in Table 3 and with *δ*_*V*_ = 1*/*180, leaving *E* and *λ*_*V*_ free.

**Table 3:** Fixed parameters values of system in Equation (1). The parameters corresponding to vaccination are established in each scenario studied. See Table 2 and Appendix A for more details.

| Parameter | Value |
| --- | --- |
| $\beta_S$ | 0.363 282 |
| $\beta_A$ | 0.251 521 |
| $\alpha_S$ | 0.092 506 9 |
| $\alpha_A$ | 0.167 504 |
| $\delta_E$ | 0.196 078 |
| $\delta_R$ | 0.002 739 73 |
| $\mu$ | 0.000 039 138 9 |
| $\theta$ | 0.11 |
| $p$ | 0.1213 |

**Figure 4:**
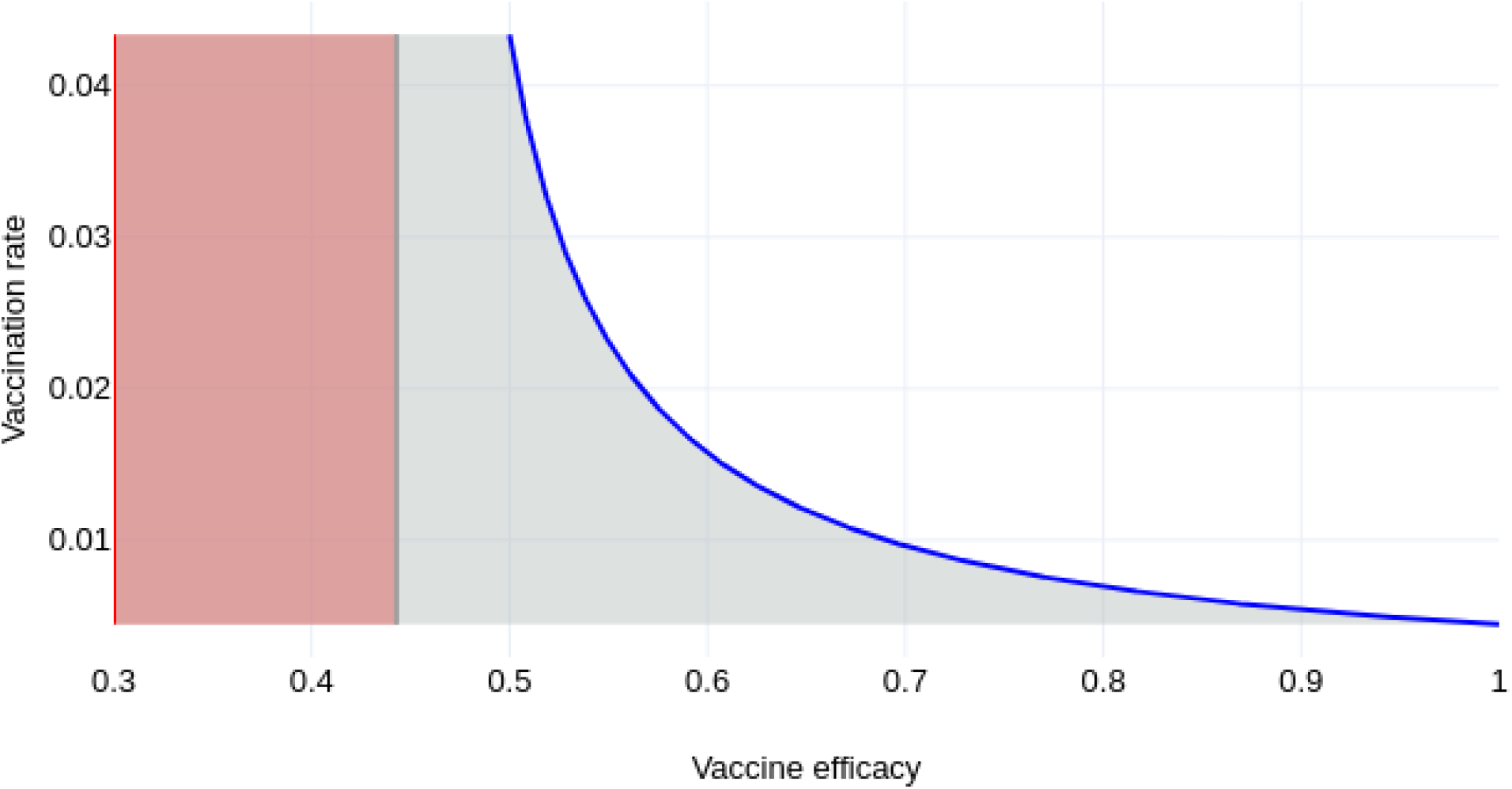
Vaccine efficacy versus vaccination rate feasibility. In the gray shaded region *R*_*V*_ *>* 1 and in the white region *R*_*V*_ *<* 1. Note that, for our scenario, with a 50 percent vaccine efficacy and an adequate vaccination rate, it is possible to reduce the *R*_*V*_ value below one. The orange region is unfeasible.

### 3.1. Baseline vaccination rate formulation

Vaccination policies to reach a given coverage of a certain percentage of the population in a given period is of great importance. In this sense, we refer to this vaccination constant rate as the base vaccination rate, denoted by *λ*_*V base*_.

Let *W* (*t*) be the normalized unvaccinated population at time *t*, we consider that at *t* = 0 no person has been vaccinated, which implies *W* (0) = 1. Then, by assuming that we vaccinate individuals at a constant rate *λ*_*V base*_ proportional to the actual population, we have that *W* (*t*) satisfies the equation

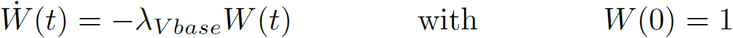

or 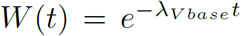. It implies that the number of vaccinated individuals at time *t* is given by 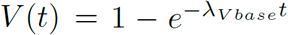. Then, if we look to reach a coverage *x*_*coverage*_ at a horizon time *T*, it follows that *λ*_*V base*_ satisfies the equation

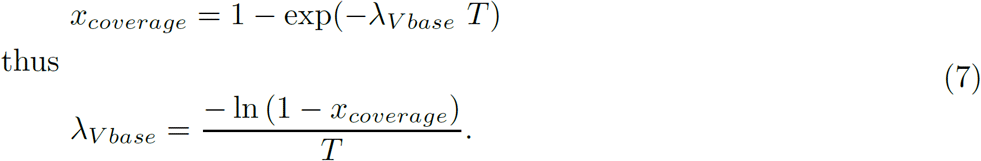

Observe that in the calculation of *λ*_*V base*_, it is considered all the population to be vaccinated. Vaccination is not applied to infective symptomatic individuals. Therefore, Equation (7) represents an approximation of the vaccination scheme at constant rate.

Figure 5, shows the contour curves for *R*_0_ considering it as a function of the efficacy of the vaccine (*E*) and of the vaccination rate (*δ*_*V*_), considering an immunity period induced by the vaccine of half year. The blue line, correspond to the values of *λ*_*V base*_ and we can see that with this vaccination rate, no matter how effective the vaccine is, it is not possible to reduce the value of *R*_*V*_ below one. Purple lines show a scenario in which it is possible to reduce the *R*_*V*_ value below one, considering a vaccine efficacy of 0.8 and a vaccination rate of 0.7. The figure shows plausible combinations of *ϵ* and *λ*_*V*_ values in order to reduce the value of *R*_*V*_ below one.

**Figure 5:**
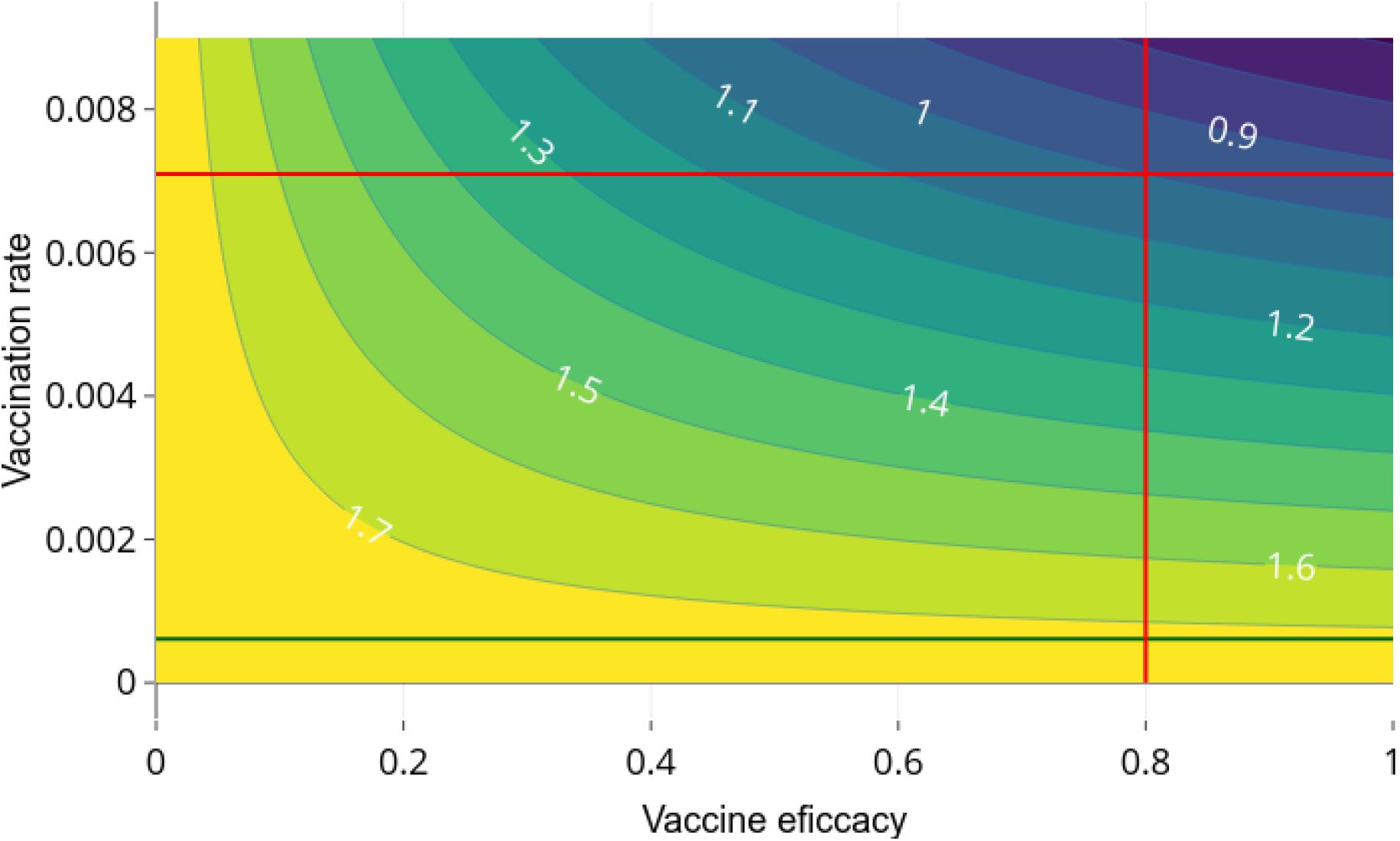
Contour plot of *R*_*V*_ as a function of *ϵ* and *λ*_*V*_ and vaccine-induced immunity average time of half year. Dark green line represents the value of *λ*_*V base*_ = 0.000 611, corresponding to a coverage *x*_*coverage*_ = 0.2 and a horizon time *T* = 365 days. Red lines show a scenario in which it is possible to reduce the *R*_*V*_ value below one, considering a vaccine efficacy of 0.8 and a vaccination rate of 0.7.

Note that a vaccine efficacy of 50 % or more is required so that, with an adequate vaccination rate, the *R*_*V*_ value can be reduced below one.

In the next section, the optimal control theory will be applied to propose optimal vaccination dynamics that minimize the number of cases of symptomatic infection and deaths due to the disease.

## 4. Optimal Vaccination policies

According to dynamics in Equations (1) and (2), we modulate the vaccination rate by a time-dependent control signal *u*_*V*_ (*t*) to achieve an imposed vaccine coverage. That is, according to the components *S, V, X* of Equations (1) and (2), we modulate the vaccination rate *λ*_*V*_ by an additive control *u*_*V*_ (*t*). Thus, we modify components equations related to *S, V, X* as

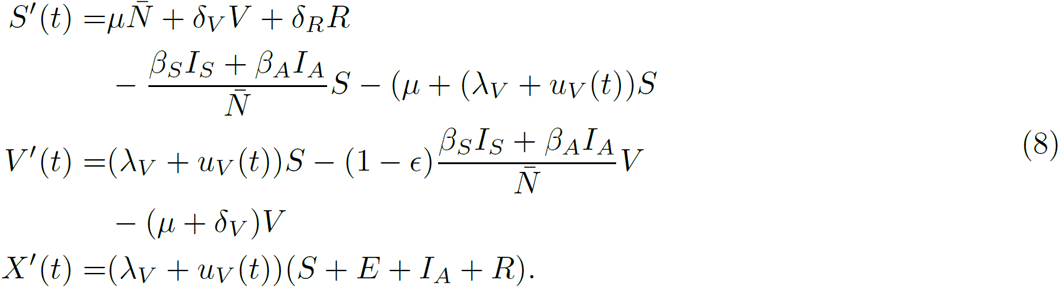

To assure the solution of our controlled model we consider the functional space

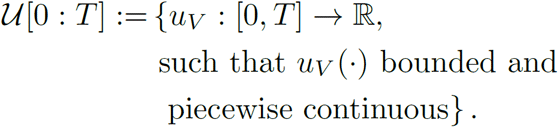

Let *x*(*t*) := (*S, E, I*_*S*_, *I*_*A*_, *R, D, V, X*)^T^(*t*) and control signal *u*_*V*_ (·) *∈𝒰* [0, *T*]. Following the guidelines of WHO-SAGE modeling questions [23], we quantify the burden of COVID-19, according to the Disability-Adjusted Life Year (DALY) unit. Thus adapting the WHO definition of DALY [24], we optimize the number of years of life lost with a vaccination policy. In other words, we calculate a minimum of the penalization functional

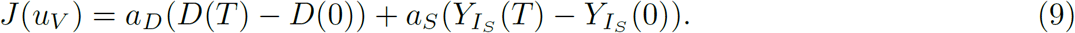

Here, *a*_*S*_ and *a*_*D*_ are parameters related to the definition of the unit of the Years of Life Lost (YLL) due to premature mortality and the Years Lost due to Disability (YLD). We estimate *a*_*D*_ as the mean life expectancy at the age of death, and according to Mexico City+Mexico state data, we handle *a*_*D*_ = 7.5 years. Parameter *a*_*S*_ is the product of a disability weight (DW) and the average duration of cases until remission or death in years, that is, 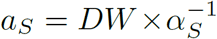. Here we postulate the disability weight as the arithmetic average of disability weight regarding comorbidities reported in [26]. Thus, our simulations employ *a*_*S*_ = 0.008 418 473 years. Thus, functional *J* penalizes the pandemic burden—in Years of Life Lost—due to mortality or disability. We display in Table 4 parameters regarding the optimal controlled model.

**Table 4:** Parameters regarding controlled version model (OCP) in Equation (12).

| Symbol | Description | Value | Ref |
| --- | --- | --- | --- |
| $a_D$ | Penalization weight due to premature mortality (YLL) and estimated from CDMX data | 7.5 years | [24, 30] |
| $a_S$ | Penalization weight due to disability (YLD) | 0.008 418 473 years | [26] |
| $x_{coverage}$ | Covering constraint at time horizon $T$ | | [23] |
| $\kappa$ | Hospitalization rate | 0.05 | Estimated |
| $B$ | Health service capacity in number of beds normalized by the whole population $N$ | 9500 | |

Since we aim to simulate vaccination policies contra factual scenarios following the SAGE modeling guidelines reported in [23], we impose the vaccination counter state’s horizon time condition *X*(*T*)

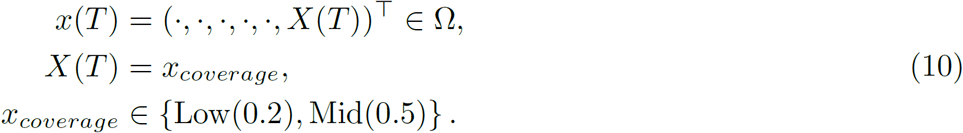

Thus, given the time horizon *T*, we set the last fraction of vaccinated population corresponds to 20% or 50%, and the rest of final states are free. We also impose the path constraint

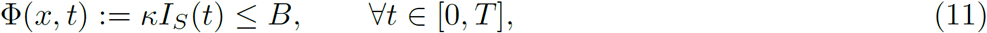

to ensure that critical symptomatic cases will not overload healthcare services. Here *κ* denotes hospitalization rate, and *B* is the load capacity of a health system. We illustrate the main ideas of the above discussion in Figure 6.

**Figure 6:**
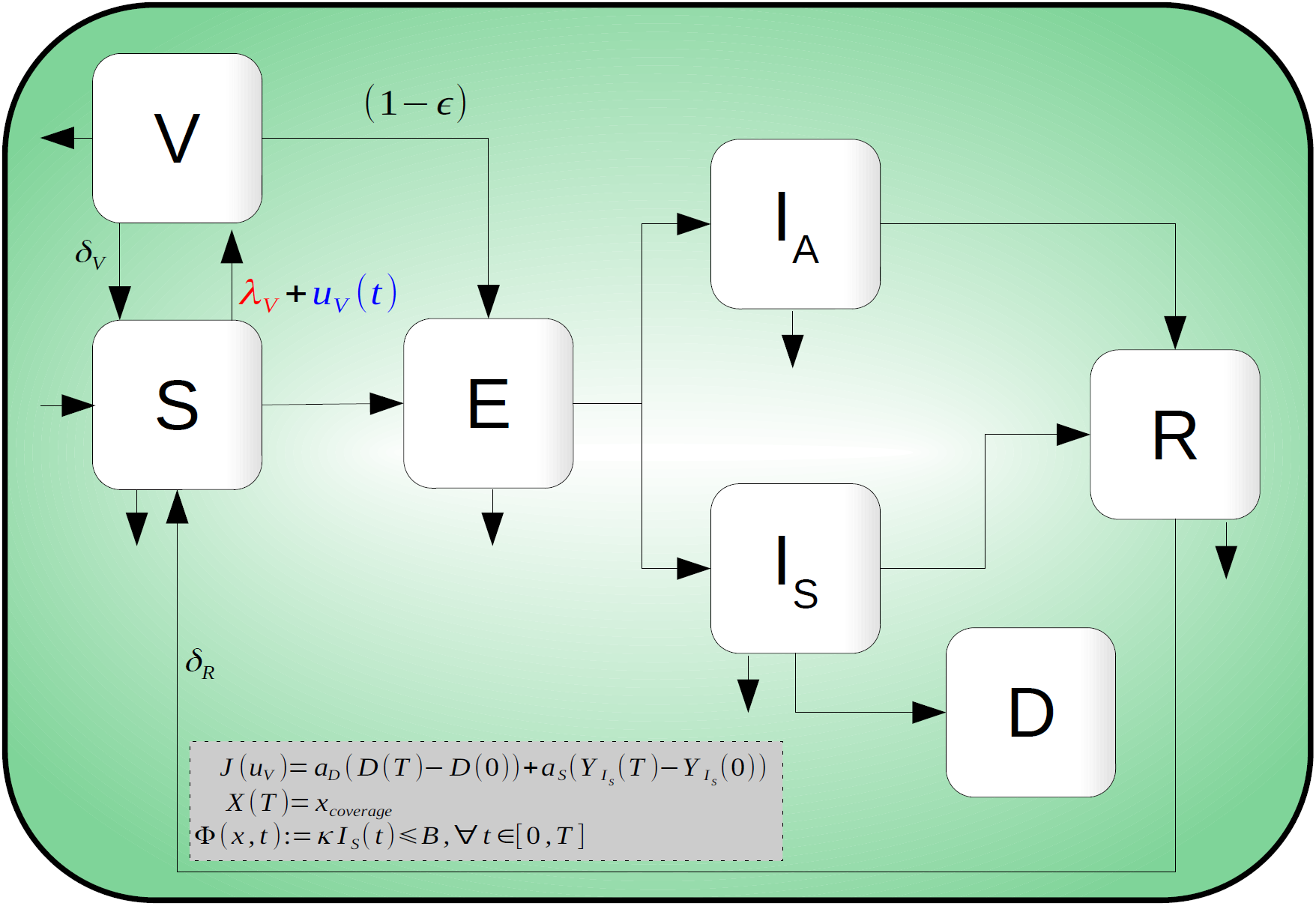
Compartmental diagram of COVID-19 transmission dynamics that includes optimal vaccination dynamics, penalization and a path constraint.

Given a fixed time horizon and vaccine with efficacy *ϵ*, we estimate the constant vaccination rate *λ*_*V*_ as the solution of Equation (7). That is, *λ*_*V*_ estimates the constant rate to cover—with a vaccine dose per individual—population fraction *x*_*coverage*_ in time horizon *T*. Thus, according to this vaccination rate, we postulate a policy *u*_*V*_ that modulates vaccination rate according to *λ*_*V*_ as a baseline.

Then, optimal vaccination amplifies or attenuates this estimated baseline *λ*_*V*_ in an interval 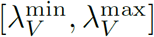 to optimize functional *J* (·)—minimizing symptomatic, death reported cases and optimizing resources.

We aim to minimize the cost functional (9)—over an appropriated space—subject to the dynamics in Equations (1) and (8), boundary conditions related to (10), and path constrain (11). That is, we seek for vaccination policies *u*_*V*_ (·), that solves the following optimal control problem (OCP)

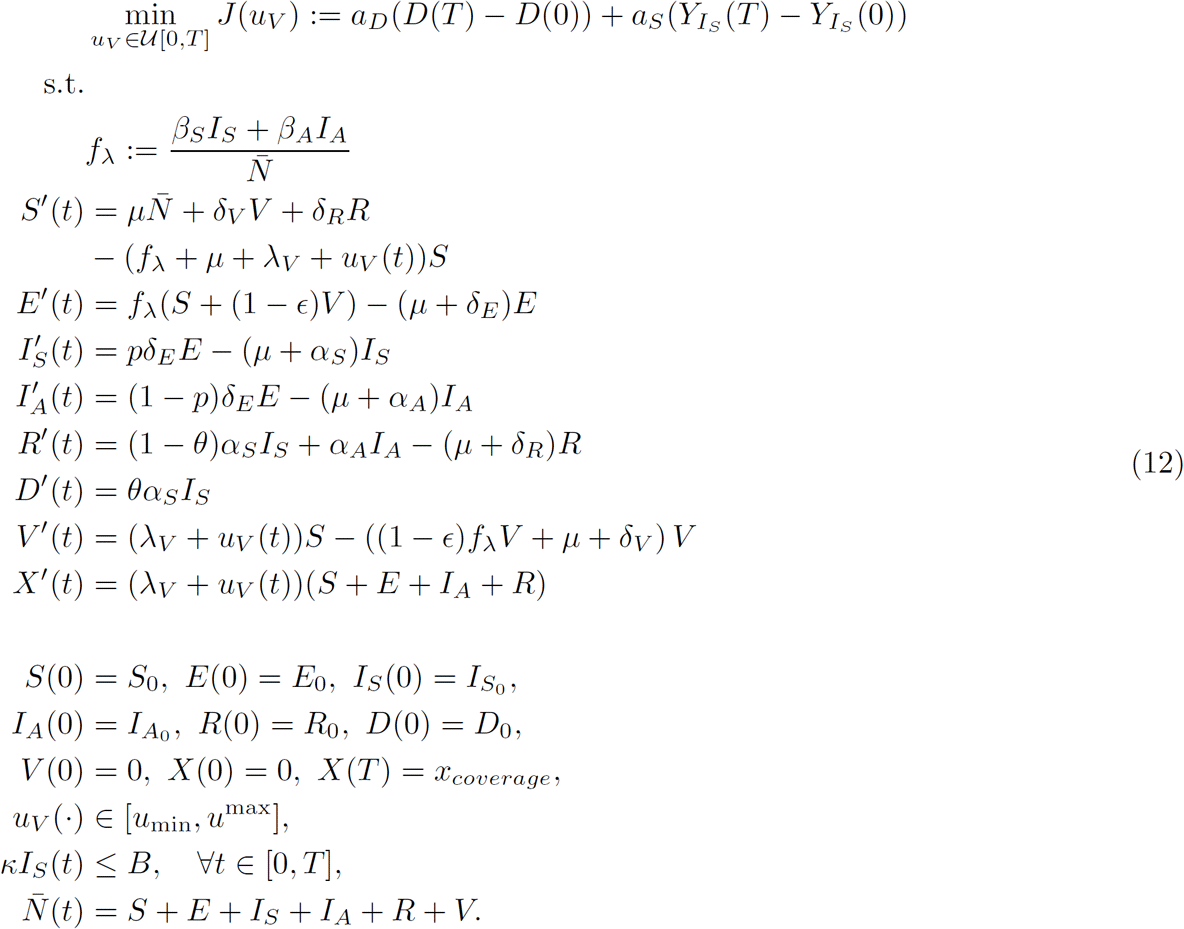

Table 4 encloses a parameter description regarding this controlled version.

Existence of solution to our (OCP) in Equation (12) drops in the theory developed by Francis Clark [see e.g. 27, Thm. 23.11]. Since our aim is the simulation of hypothetical scenarios, we omit here a rigorus proof, instead we refer interested readers to [28, 29] and the reference there in.

## 5. Numerical experiments

### 5.1. Methodology

We apply the so-called transcript method to solve our (OCP). This schemes’ main idea consists of transforming the underlying problem of optimizing functional governed by a differential equation into a finite-dimensional optimization problem with restrictions. To fix ideas, let *x, u* denote state and control and consider the optimal control problem

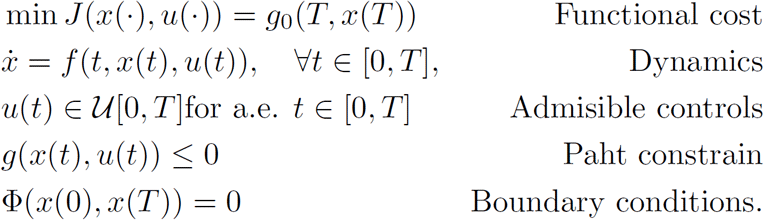

Then, transcription methods transform this infinite-dimensional optimization problem into a finite dimension problem (NLP) via discretization of dynamics, state, and control. For example, if we employ the Euler method with a discretization of *N* constant steps with size *h*, then we can solve

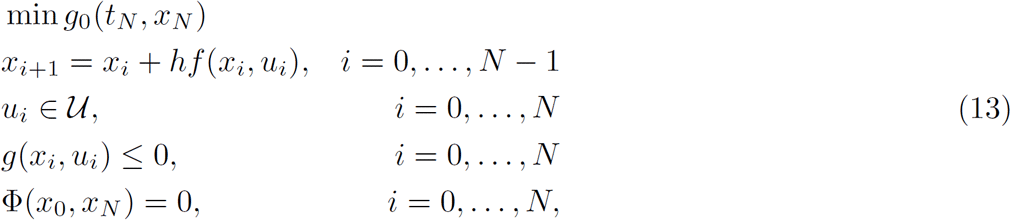

where *x*_*i*_ *≈ x*(*t*_*i*_), *u*_*i*_ *≈ u*(*t*_*i*_) in the grid

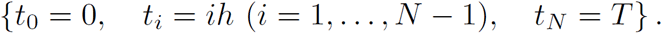

Let *Y* = {*x*_0_, *…, x*_*N*_, *u*_0_, *… u*_*N*_}. Thus Equation (13) defines a nonlinear programming problem on the discretized state and control variables of the form

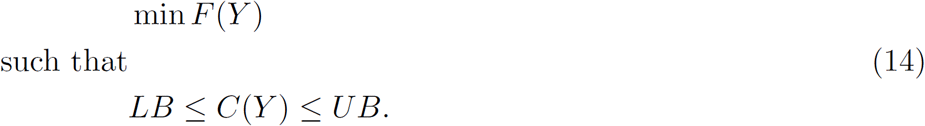

The numerical analysis and design of transcript methods is a well established and active research numerical field. There is a baste literature about robust methods and resently it apperars implementations in vogue languages like Julia [31, 32], Python [33], Matlab [34], and others. We refer the reader to [35, 36] for a more systematic discussion.

Our simulations rely on the `Bocop` package [37, 38] to solve our (OCP). Bocop is part of the development of the INRIA-Saclay initiative for open source optimal control toolbox and supported by the team Commands. BOCOP solves the NLP problem in Equation (14) by the well known software Ipopt and using sparse exact derivatives computed by ADOL-C.

We provide in [39] a GitHub repository with all regarding R and Bocop sources for the sake of reproductivity. This repository also encloses data sources and python code to reproduce all reported figures.

### 5.2. Simulation of hypothetical scenarios

We follow the guidelines reported by the WHO Strategic Advisory Group of Experts (SAGE) on Immunization Working Group on COVID-19 Vaccines modeling questions presented in [23]. According to this SAGE’s document, we simulate scenarios to illustrate vaccination policies’ response with a preventive vaccine. We aim to contrast the impact of the burden of COVID-19 mitigation regarding

(**SCN**-1) Optimal versus constant vaccination policies

(**SCN**-2) Vaccine efficacy

(**SCN**-3) Induced vaccine immunity

(**SCN**-4) Natural immunity

We consider vaccine profiles—efficacy and induced vaccine immunity in concordance with the expected but still unconfirmed data—from the firms Cansino-Biologics, Astra Zeneca, and Pfizer. Further, since reinfection and induced vaccine immunity parameters remain unavailable, we see pertinent explore the effect of plausible settings.

#### Remark 1.

We mean an optimal vaccination policy as the number of doses per unit time described by

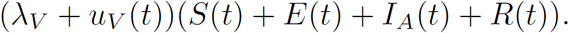

Counterfactual scenarios implies *u*_*V*_ (*t*) = 0. Without vaccination scenarios we means *λ*_*V*_ = *u*_*V*_ (*t*) = 0.

Table 5 encloses a brief description and parameter values regarding each scenario. The reader can also access the web `Chart Studio Graph` of each figure regarding data and `plotly` [40] visual representation.

**Table 5:**
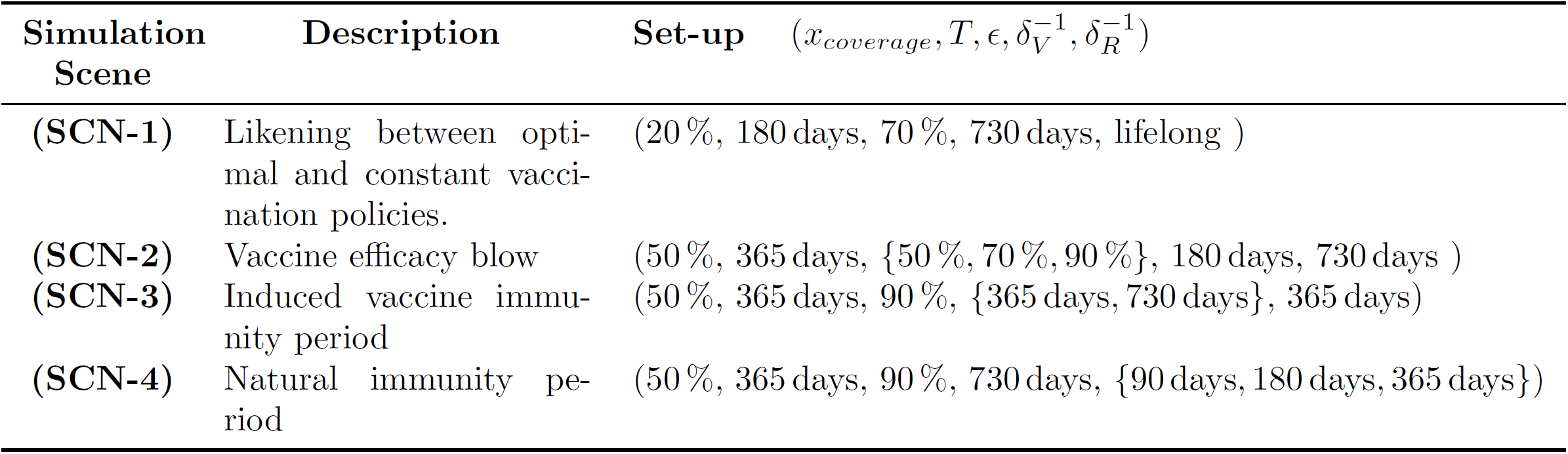
Setup parameters for counterfactual and response scenarios. See Table 3 for the rest of parameters.

| Simulation Scene | Description | Set-up $(x_{coverage}, T, \epsilon, \delta_V^{-1}, \delta_R^{-1})$ |
| --- | --- | --- |
| (SCN-1) | Likening between optimal and constant vaccination policies. | (20 %, 180 days, 70 %, 730 days, lifelong ) |
| (SCN-2) | Vaccine efficacy blow | (50 %, 365 days, {50 %, 70 %, 90 %}, 180 days, 730 days ) |
| (SCN-3) | Induced vaccine immunity period | (50 %, 365 days, 90 %, {365 days, 730 days}, 365 days) |
| (SCN-4) | Natural immunity period | (50 %, 365 days, 90 %, 730 days, {90 days, 180 days, 365 days}) |

To perform the simulations corresponding to the scenarios presented in Table 5, we fix the parameter values as in Table 6.

**Table 6:**
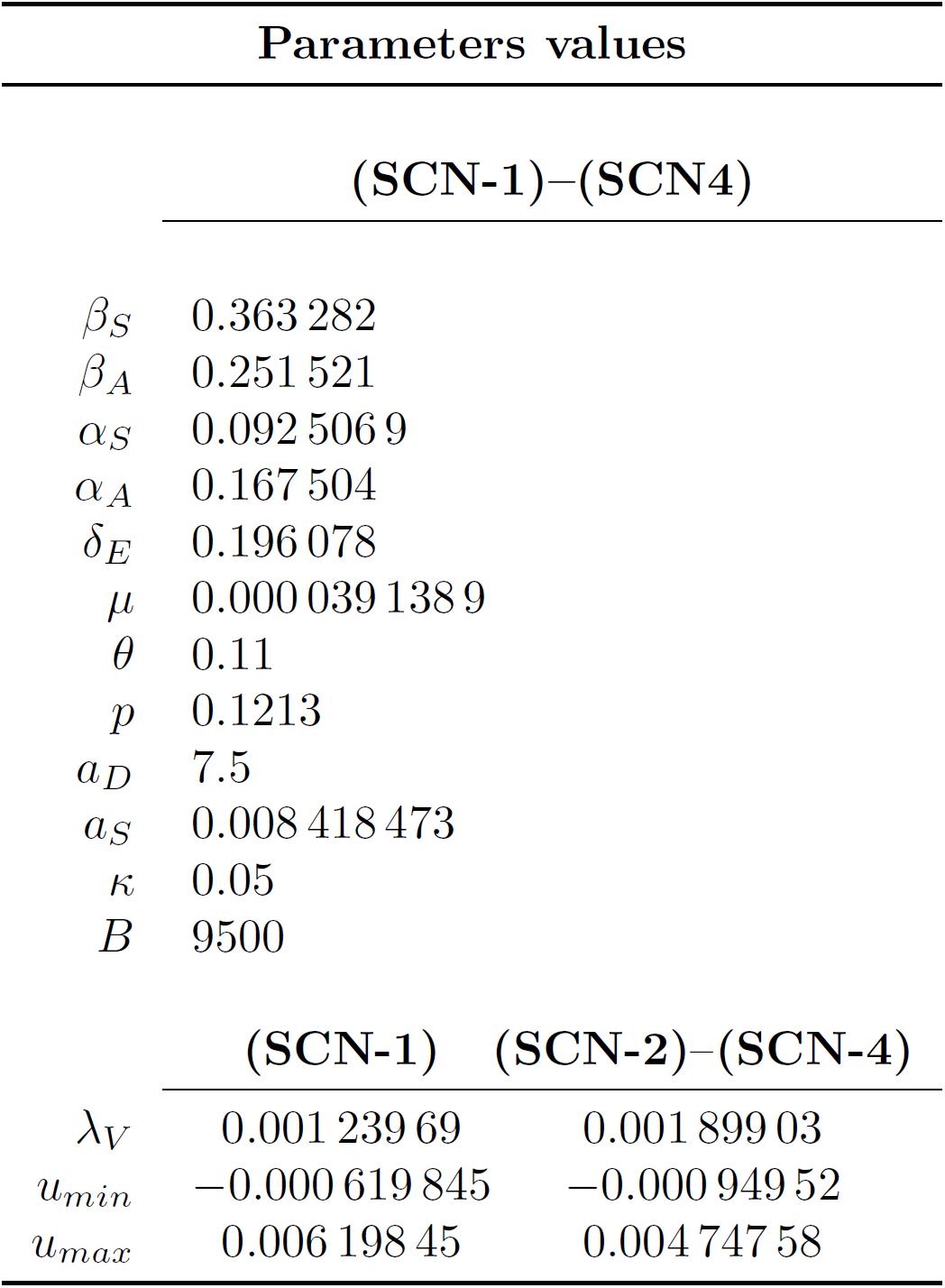
Fixed parameters values of system in Equation (12).

#### Optimal Versus Constant Vaccination Policies: (SCN-1)

To fix ideas, we display in Figures 7 and 8 the counterfactual scenario regarding no intervention, constant vaccination policy (CP), and optimal vaccination policy (OP) with a vaccine profile of efficacy ∊ = 70 %, and induced immunity 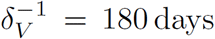 days and over a campaign for 20 % of coverage at 180 days. Figure 7 suggests that the OP improves CP vaccination policy response according to the burden disease due to mortality, morbidity, and coverage time. Figure 8 confirms this improvement by comparing symptomatic reported cases, saving lives, and the disease dynamics without vaccination. Although both campaigns use the same number of vaccine doses and the same vaccine profile, we observe that OP implies fewer deaths and symptomatic cases.

**Figure 7:**
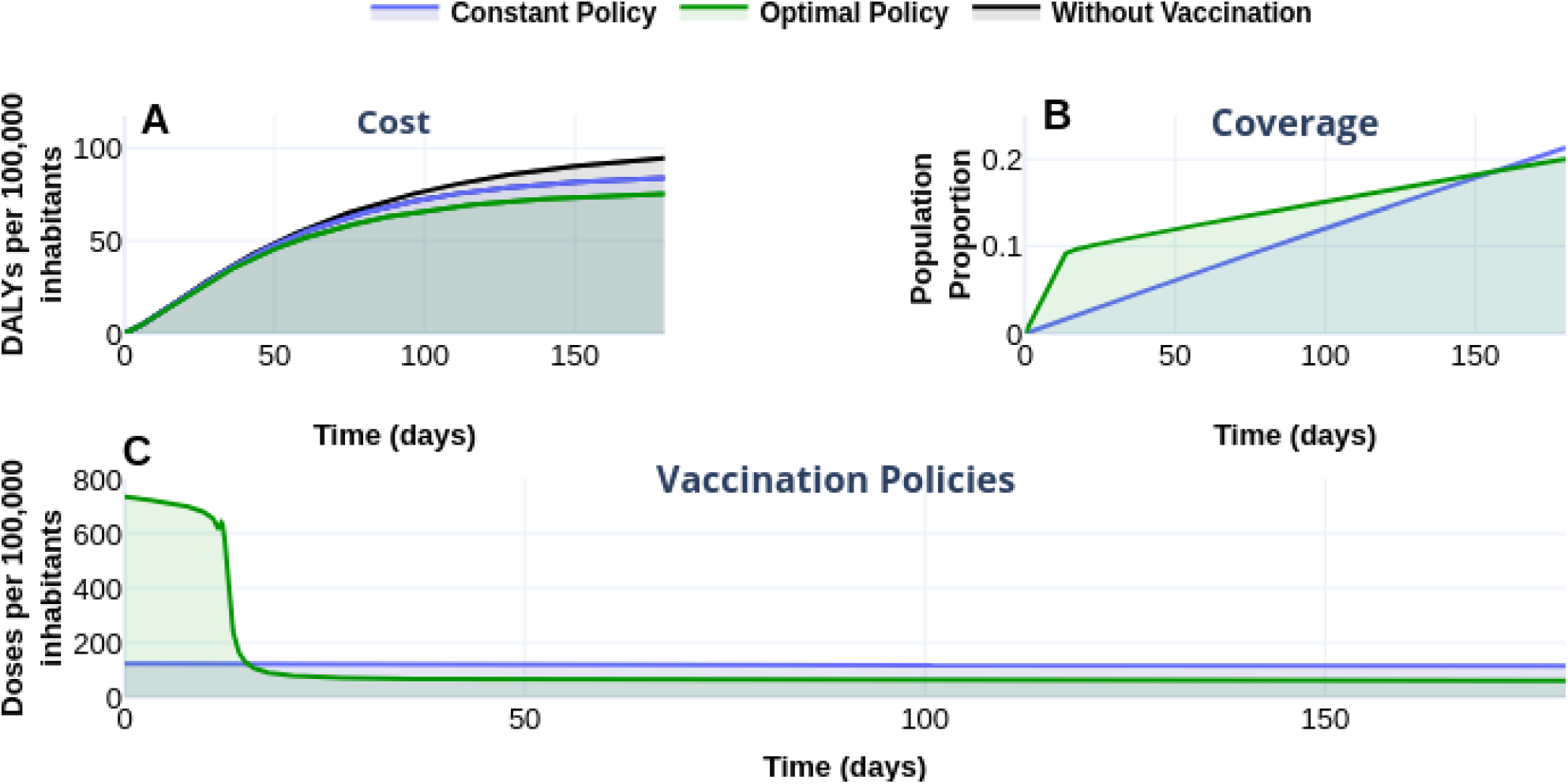
Effect of the vaccination policy on the burden COVID-19. (A) vaccination policies’ response regarding constant (*λ*_*V*_) and optimal (*λ*_*V*_ +*u*_*V*_ (*t*)) vaccination rates in the burden of COVID-19 quantified in DALYs. Blue translucent color corresponds to policies with constant vaccination rate 0.001 239 69. Green tone denotes the vaccination optimal rate. For counterfactual reference, black line-gray shade represents the burden of COVID-19 without vaccination. (B) evolution of the vaccination covering according to each policy. Optimal policy requires less time to reach 20 percent of the vaccinated population. (C) vaccination schedule for each vaccination policy. See https://plotly.com/sauldiazinfante/131/ for plotly visualization and data.

**Figure 8:**
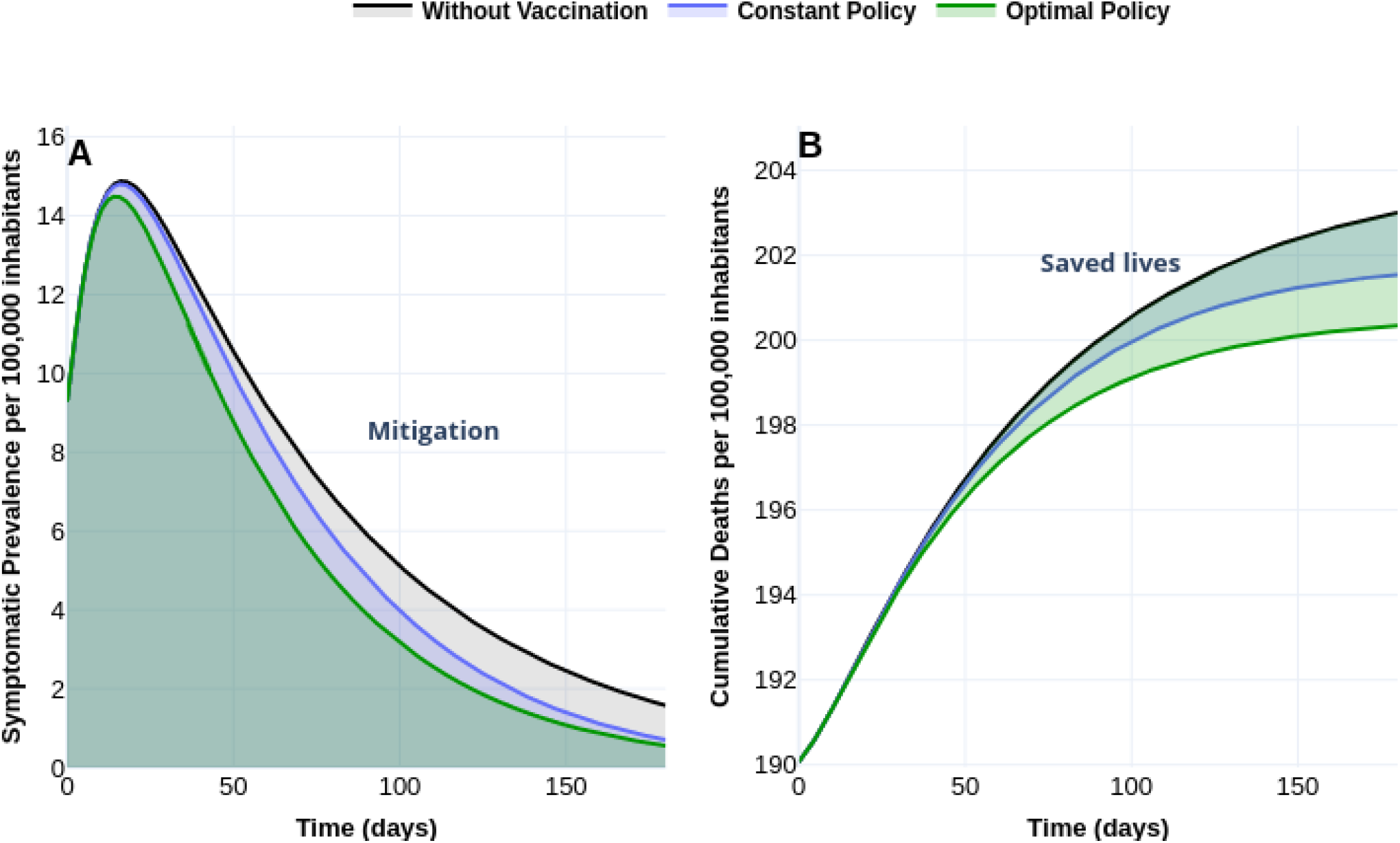
Effect of the vaccination policy on outbreak evolution. (A) Optimal vaccination policy reaches a better response in mitigating symptomatic cases than a policy with a constant vaccination rate. (B) Shaded translucent regions denote the number of saved lives per 100 000 inhabitants according to constant (blue) or optimal vaccination rates. Since the areas overlap with a green tone, we note that optimal vaccination policy improves the number of saved lives. Data and web visualization in https://plotly.com/sauldiazinfante/135/.

Figure 7, shows a scenario where *R*_0_ *>* 1. Although *R*_*V*_ remains below but close to *R*_0_, vaccination reproductive number *R*_*V*_, could explain vaccination response with constant policies according to the mitigation factor

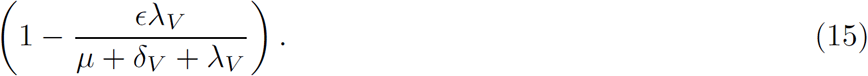

That is, disease mitigation is strongly related to vaccine efficiency *E* and vaccination rate *λ*_*V*_. Further, given a dynamic with not vaccine intervention and *R*_0_ *>* 1, *R*_*V*_ suggests a minimal vaccination rate to drive this dynamic to the disease-free state but subject to vaccines with particular efficacy.

#### Vaccine Efficacy (SCN-2)

In the author’s words of press releases

> “Pfizer says early analysis shows its Covid-19 vaccine is more than 90 % effective [41].”
>
> “Russia says its Sputnik V COVID-19 vaccine is 92 % effective [42].”
>
> “Moderna’s coronavirus vaccine is 94.5 % effective, according to company data [43].”

This is a game changer fact—FDA would accept a vaccine of 50 %. Following this idea, Figures 9 and 10 display the response of the optimal vaccination policy according to three vaccines with different efficacy. Figure 9-A displays the burden COVID-19 in DALYs for vaccines with efficacy of 50 %, 70 %, 90 %. According with a time horizon of 1 year and coverage of 50 %, Figure 10 displays an improvement of at least three times in the prevalence of symptomatic cases and saved lives concerning the uncontrolled outbreak. Figure 10 reflects this effect in the prevalence of symptomatic cases (A) and in the number of saved lives shaded by the translucent green color (B).

**Figure 9:**
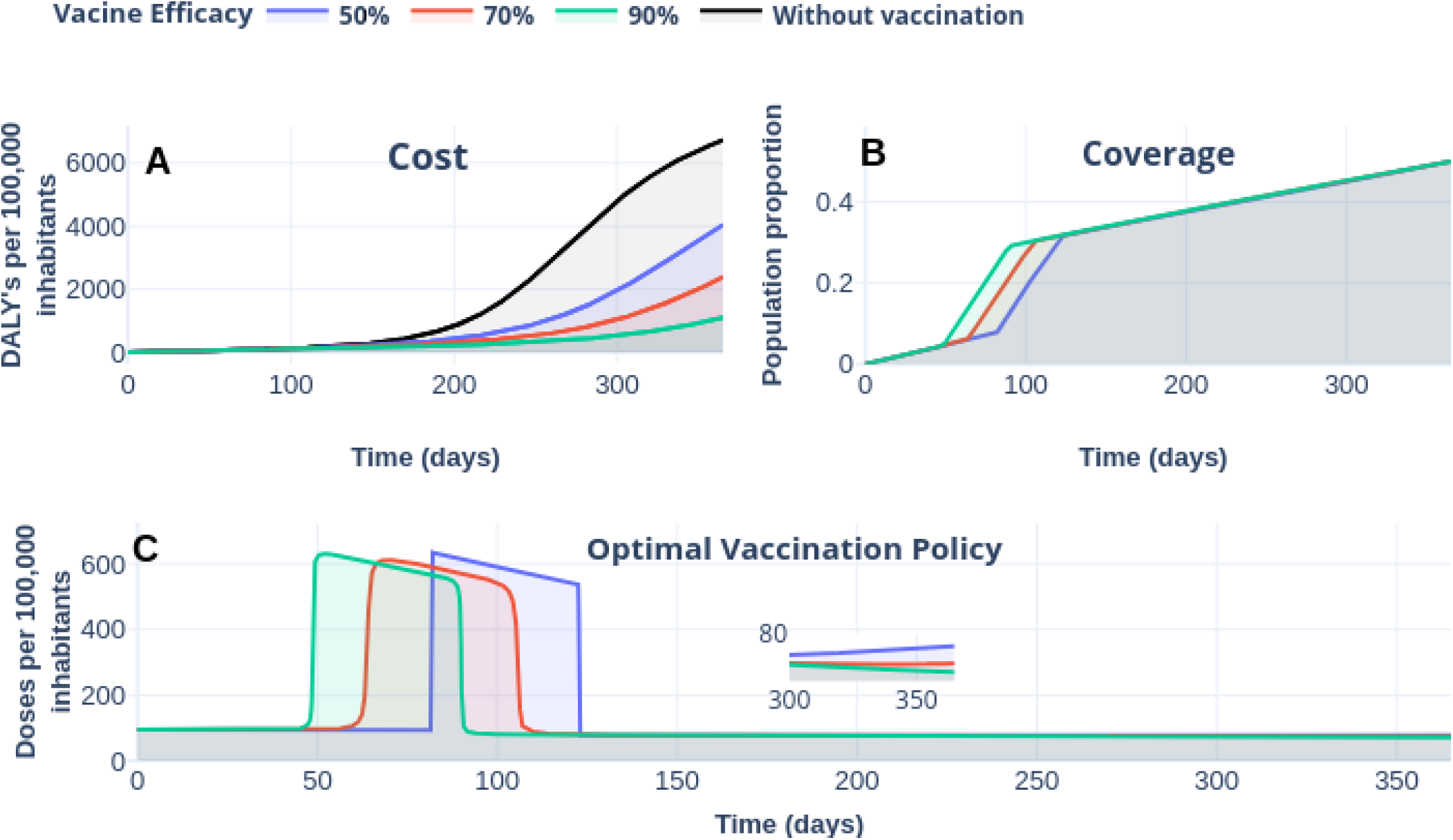
The response of COVID-19 burden on vaccine efficacy. (A) COVID-19 burden response quantified in DALYs per 100 000 inhabitants to vaccines with efficacy of 50 % (blue), 70 % and 90 %(red). (B) Coverage evolution to reach 50 % of the total population vaccinated. (C) Optimal vaccination doses schedule according to the different efficacies. See https://plotly.com/sauldiazinfante/85/ for visualization and data.

**Figure 10:**
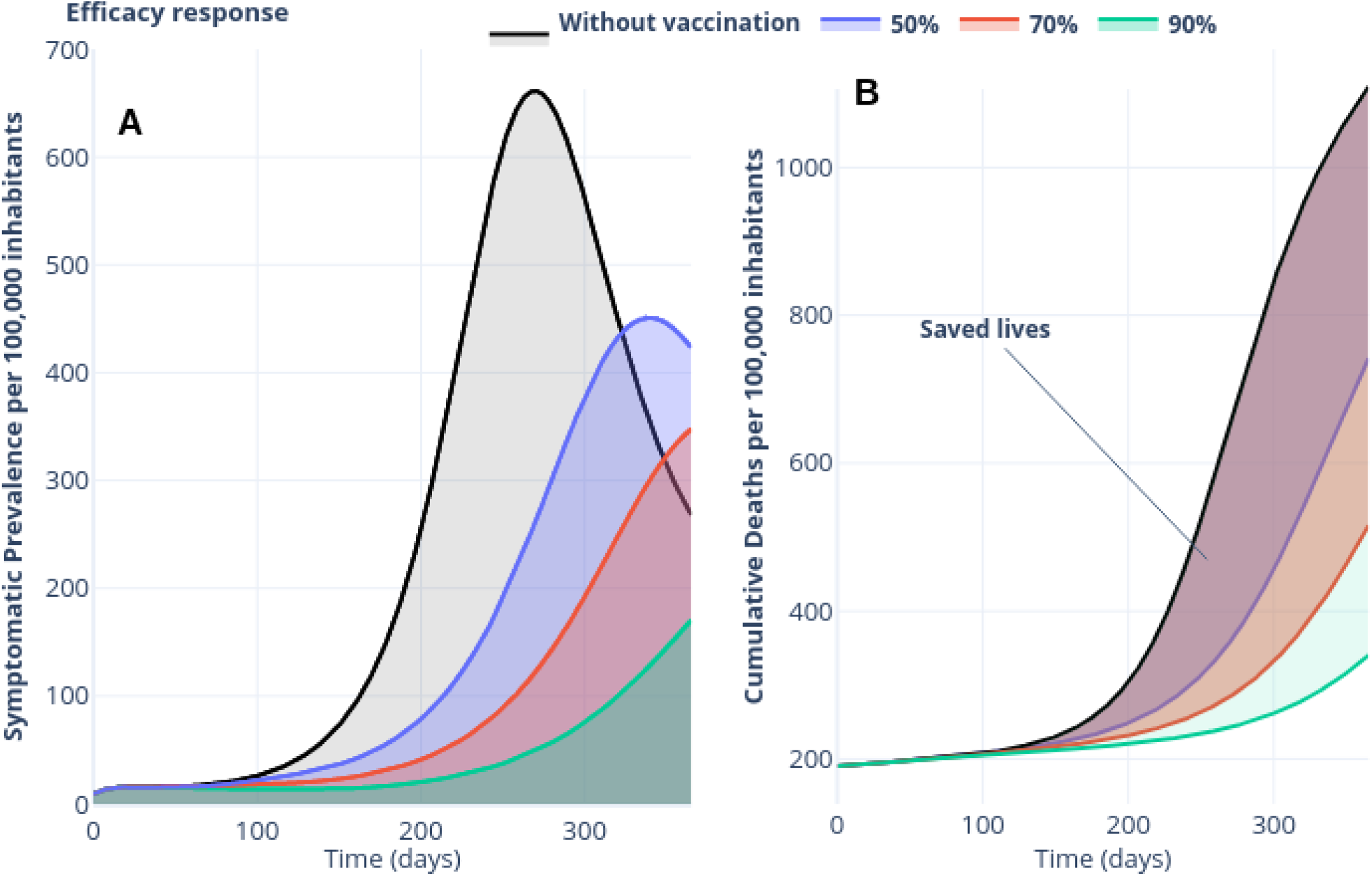
(A) Effect of vaccine-efficacy of 50 % (blue), 70 % and 90 % (red) on prevalence of symptomatic cases per 100 000 inhabitants. (B) Effect of vaccine-efficacy on the number of saved lives. See https://plotly.com/sauldiazinfante/100/ for data and visualization.

#### Vaccine-induced immunity (SCN-3)

Vaccine response is also strongly related to its induced immunity —parameter that remains poorly understood [44]. Here we contrast two vaccines with different induced immunity. Let denote by *vax*_1_, *vax*_2_ vaccines with an induced-immunity capacity of one and two years, respectively, and common efficacy of 70 %. Consider a vaccine camping of time horizon of one year and 50 % coverage. Taking the same dynamics parameters, that is initial conditions, and base line parameters as in Table 3 we explore a contra factual scenario with an uncontrolled outbreak of *R*_0_ = 1.794 93 and two controlled dynamics according to vaccines *vax*_1_, *vax*_2_. Thus, according to these immunity parameters and factor defined in Equation (15), respectively, results 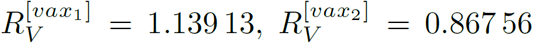 for vaccine immunities periods of one and two years. We display in Figure 11 the response of the vaccines *vax*_1_ and *vax*_2_. Since in this set time horizon is of one year, the optimal policies follows similar schedules and implies similar gains in the number of years of life lost. Despite this similarities, Figure 12 displays in panel A a dramatically gain respect to the uncontrolled outbreak—since 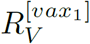 is near to one, prevalence fall-down more than five times and because 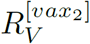 is lest that one, prevalence of symptomatic cases tends to zero with damped oscillations. Figure 12 also endorses this gain, note that saved lives is represented by shaded region with translucent and overlapped red blue colors.

**Figure 11:**
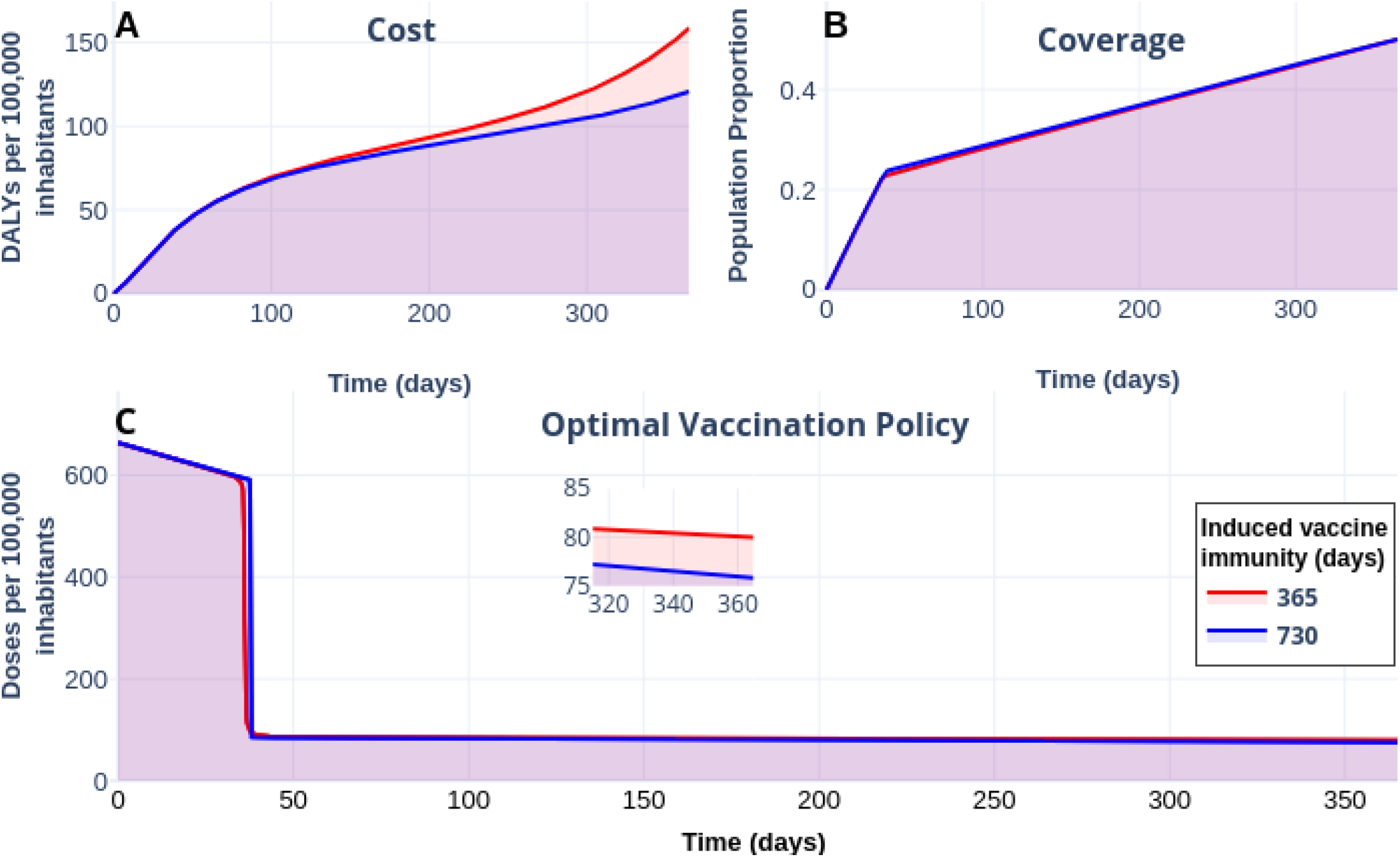
(A) Effect on the burden of COVID-19 quantified in DALYs per 100 000 inhabitants due to vaccine-induced immunity of 365 days(red) and 730 days (blue). (B) Coverage evolution to reach 50 % of the total population vaccinated. (C) Optimal vaccination doses schedule according to the different vaccine-induced immunities. Since the time horizon is 350 days, both policies follow similar profiles in coverage and schedule. Visualization and data in https://plotly.com/sauldiazinfante/111/.

**Figure 12:**
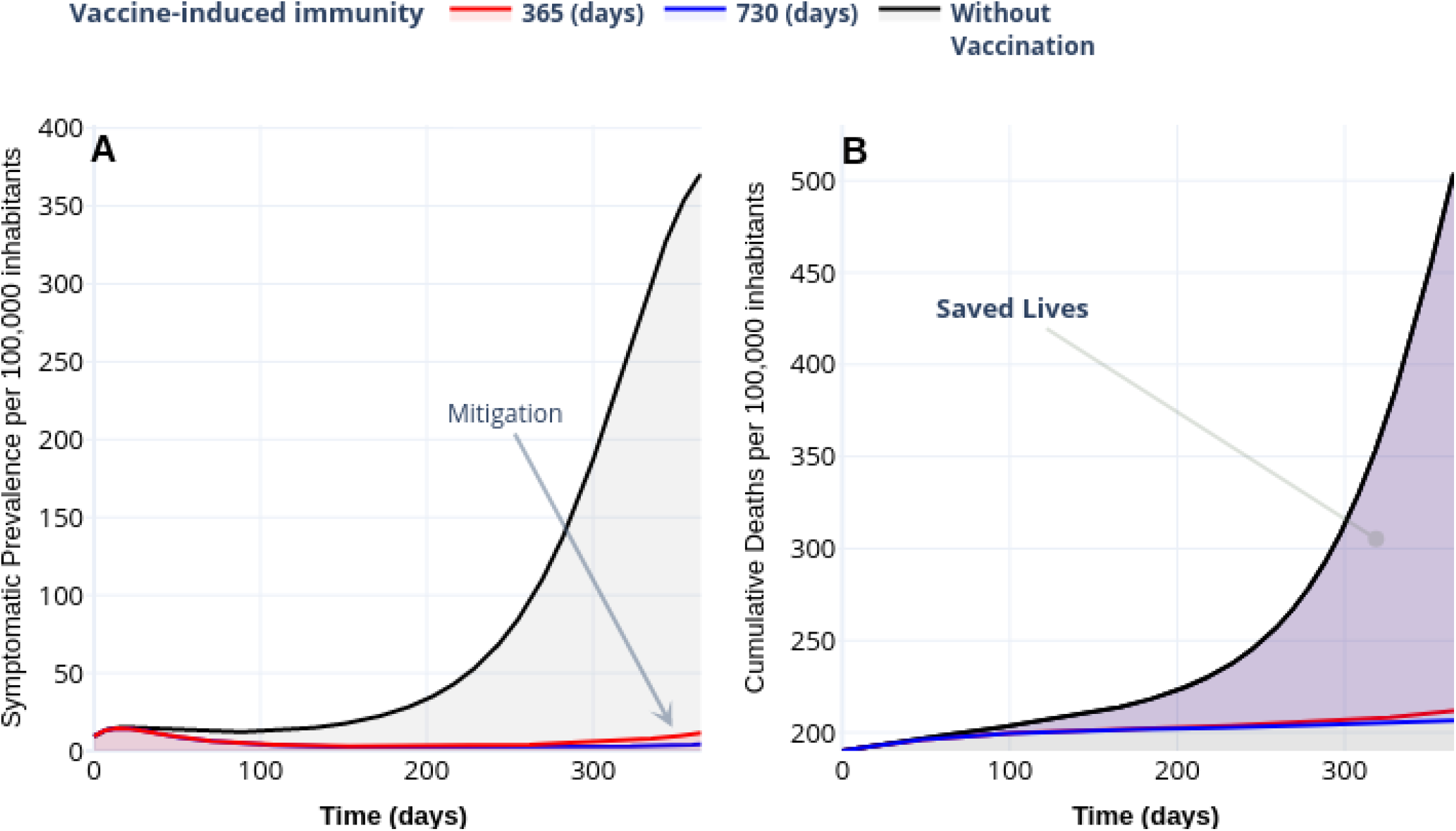
(A) Effect of vaccine-induced immunity on mitigation of symptomatic prevalence per 100 000 inhabitants. (B) Number of saved lives. Since the reproductive vaccine number for the immunity of 365 days results in 1.139 13, and 0.867 56 for 730 days, this behavior is consistent. See https://plotly.com/sauldiazinfante/123/ for data and visualization.

#### Natural Immunity Hypothesis (SCN-4)

“Second infections raise questions about long-term immunity to COVID-19 and the prospects for a vaccine”, reported Heidi Ledford in [45]. Following this line, we display in Figures 13 and 14 the vaccine’s response with 90 % efficacy and contrasting with natural immunity periods of 90 days, 180 days, 365 days. Here, the adjective “natural” denotes the immunity that an individual develops after recovering from a previous bout of COVID-19. When natural immunity lasts one year, the burden of COVID-19 fall-down until around 120 DALYs. We confirm this behavior in the prevalence of symptomatic cases and cumulative deaths, as displayed in Figure 14. When natural immunity is 365 days the gain in mitigation concerning a natural immunity of 90 days is at least and 100 times, while the number of deaths with a natural immunity of 90 days reach 845 cases per 100 000 inhabitants, in contrast, of 206 when natural immunity is 365 days. Thus, this simulation suggests that natural immunity plays a vital role in the controlled outbreak’s behavior, which is consistent with the conclusions reported in [44].

**Figure 13:**
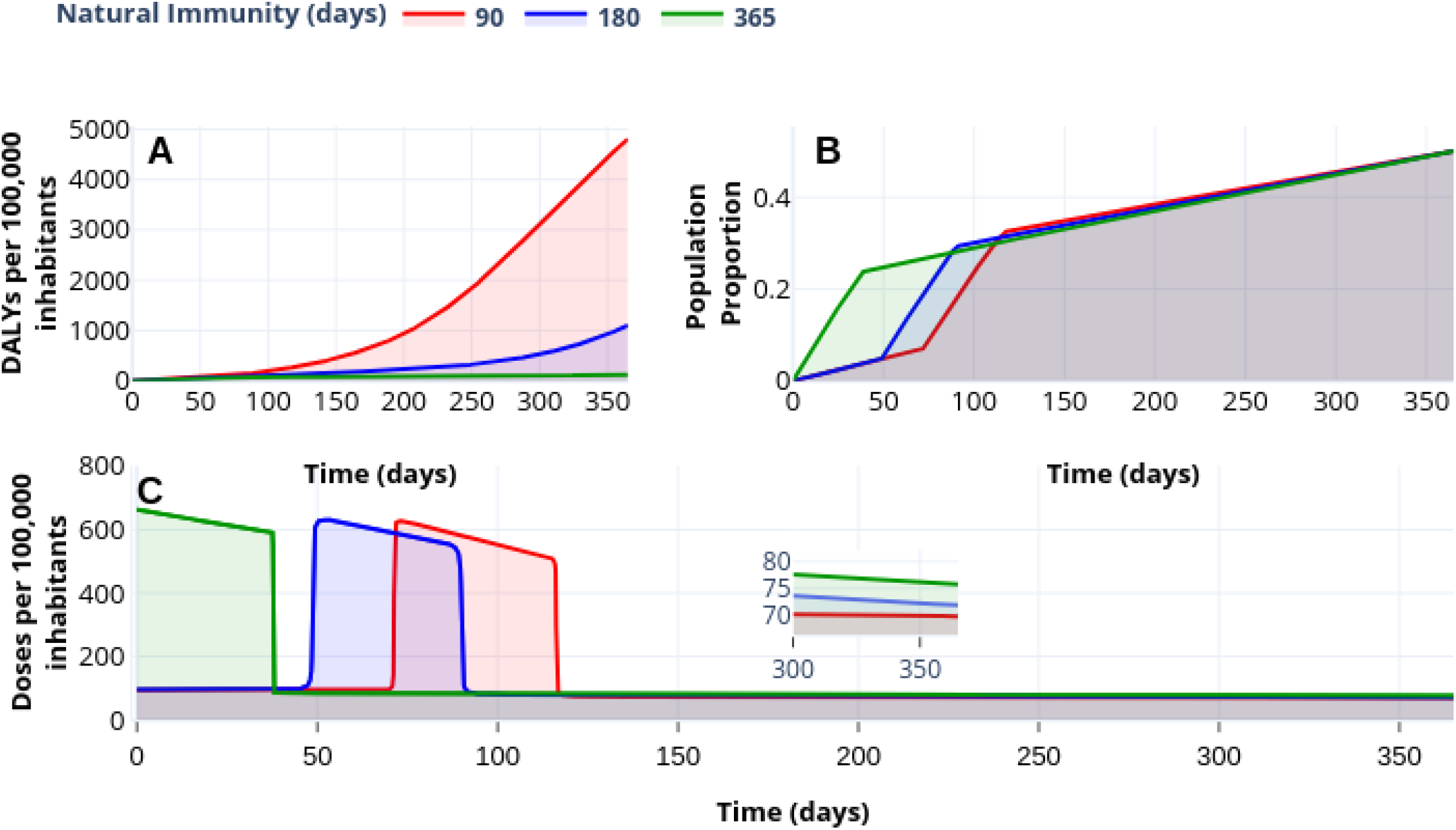
(A) Effect on the burden of COVID-19 quantified in DALYs per 100,000 inhabitants due to natural immunity of 90 days (red), 180 days (yellow) and 365 days (green). (B) Coverage evolution to reach 50 of the total population vaccinated. (C) Optimal vaccination doses schedule according to the different natural immunities. https://plotly.com/sauldiazinfante/95/

**Figure 14:**
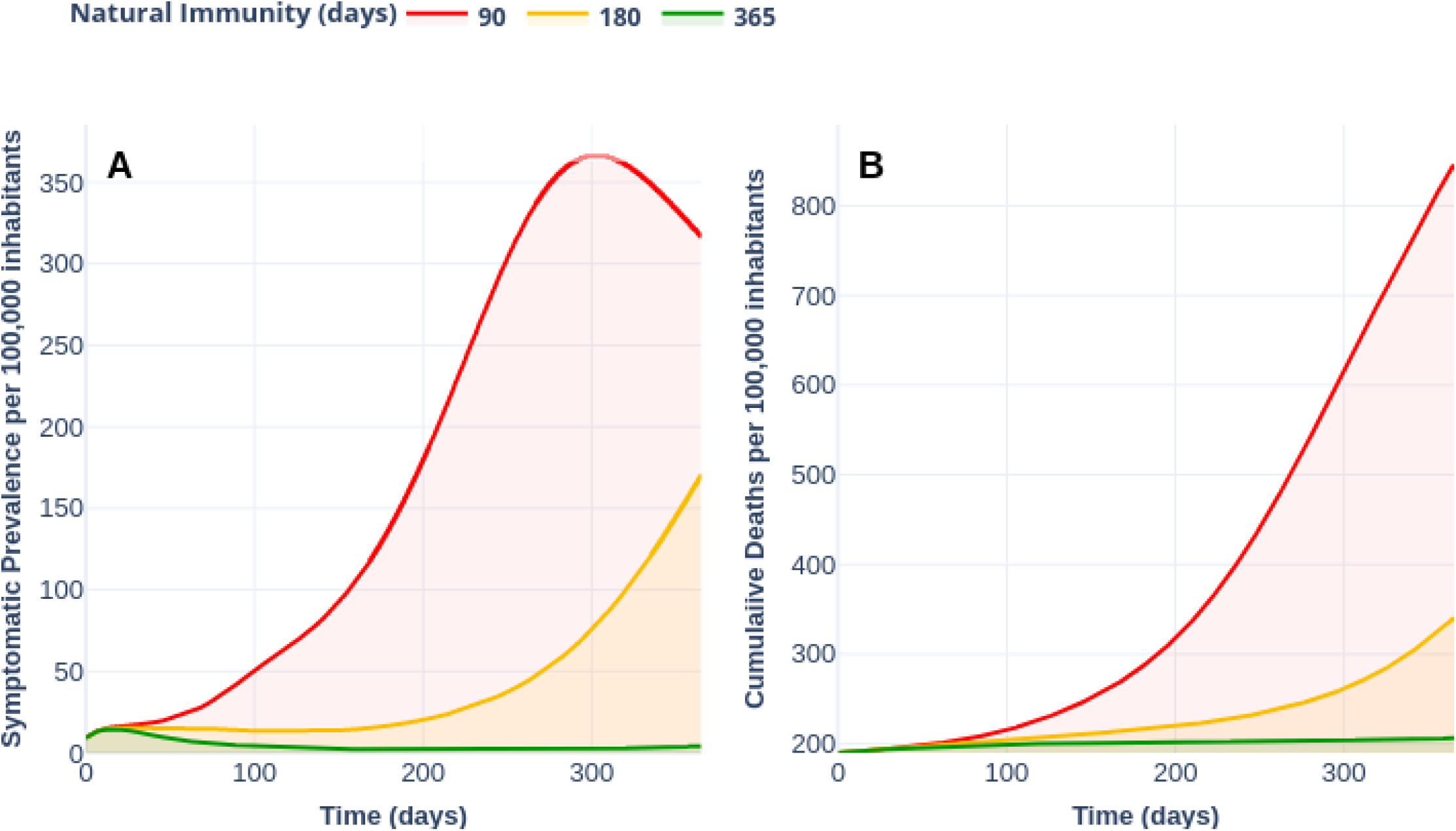
(A) Effect of immunity on mitigation of symptomatic prevalence per 100,000 inhabitants. (B) Number of saved lives. Since the reproductive vaccine number for the immunity of 365 days results in 1.139 13 and 0.867 56 for 730 days, this behavior is consistent. Plotly visualization and data in https://plotly.com/sauldiazinfante/104/.

## 6. Conclusions

At the date of writing this article, humankind lacks strategies to eradicate COVID-19. Although NPIs implemented in most countries prevent citizens from being infected, these strategies leave them susceptible—people can not develop immunity to face futures waves. Thus, vaccination becomes the primary pharmaceutical measure to recover life’s style before the pandemic. However, this vaccine has to be effective and well implemented in global vaccination programs. Thus new challenges as its distribution, stocks, politics, vaccination efforts, among others, emerge. A fair distribution and application strategy is imperative to manage the available resources, especially in developing countries.

We established an optimal control problem to design vaccination strategies where vaccination modulates dynamics susceptibility through an imperfect vaccine. We aimed to provide vaccination policies that minimize the lost life years due to disability or premature death by COVID-19, determined by cumulative deaths and cumulative incidence. Policies’ acts in the minimization of infected people’s prevalence and the number of deaths.

Our simulations suggest a better response with optimal vaccination policies than policies with a constant vaccination rate. For example, the optimal policy schedule in scenario **SCN-1** increases the number of doses in the scheme’s initial stage. This vaccination scheme improves the mitigation of the symptomatic prevalence, the incidence of deaths, and in consequence, the years of life lost quantified in DALYs.

Emerging press releases reported that Pfizer’s, Russian Sputnik V, and Moderna’s coronavirus vaccines reach efficacy over 90 % [41–43]. However, this information remains under development. Thus vaccine efficacy scenarios of 50 %, 70 %, and 90 % **(SCN-2)** illustrate the effect on optimal vaccination policies’ schedule by pointing when to intensify the number of doses. According to the time horizon of one year and coverage of 50%, our numerical experiments suggest that 90 % vaccine efficacy reduces around three times the number of deaths regarding the dynamics without vaccination. Likewise, these vaccines reach a gain of eighteen times in the years of life lost compared to the without vaccination scenario.

Our numerical experiments also illustrate vaccine-induced immunity’s relation between the reproductive vaccination number *R*_*V*_ and vaccination policies **(SCN-3)**. Considering an outbreak with a reproductive number *R*_0_ of 1.794 93, vaccine-induced immunity of 365 or 730 days implies a reduction of *R*_0_—dropping its value respectively to 1.139 130 and 0.867 56. Likewise, optimal policies linked to vaccine-induced immunities enhance symptomatic prevalence mitigation and the number of saved lives. Moreover, according to the initial number of deaths, the scenario without vaccination accumulates 503 deads compared to 211 and 206 deads of the underlying dynamics with vaccine-induced immunities.

Barbosa et al. recently report in [22] a simpler model about modeling of COVID-19 vaccination with a similar approach. Although they establish a less detailed model —they do not distinguish between symptomatic and asymptomatic infected individuals—its control problem return policies according to multi-objective policies. Their optimal policy is pragmatic but, in our opinion, not necessarily practical for large populations. Further, our model extends the result of [22] by a more detailed vaccine profile. Thus we can evaluate how vaccine efficacy, vaccine-induced, and natural immunity parameters impact the mitigation of an optimal vaccination schedule.

Perkins and España report in [20] a vaccination model with optimal control, but they approach to optimize the NPIs. The methodology presented here is similar, but aim very different. However, we want to stress the relevance of also including NPIs effects.

Since any vaccine’s efficacy will be subject to uncertainty and immunization regarding COVID-19 remains under development, policymakers need better modeling tools to design fair vaccination programs. We faced this problem by simulation.

According to DALYs definition, segregation as age, comorbidities, and other risk groups is imperative to design more realistic vaccination policies. Moreover, it is well-known that various vaccines platforms and strategies are developing in parallel, and the most recent advance is with vaccines that require two doses. From [20], we can deduce that NPIs, together with vaccination, would constitute a better description of COVID-19 control. We will direct our attention to extend this work according to the segregation and optimization of NPIs-vaccination controls.

## Data Availability

Mexico COVID-19 data is available on the Mexican government website.

https://www.gob.mx/salud/documentos/datos-abiertos-152127

## Data availability

All code is available at

https://github.com/SaulDiazInfante/NovelCovid19-ControlModelling/tree/master/UNISON-ITSON-VACCINATON-PRJ

## Authors’ contributions

The first and second authors named are leads. The second named author is the corresponding author. All other authors are listed in alphabetical order.

**Manuel Adrian Acuña-Zegarra:** Conceptualization, Methodology, Software, Validation, Formal analysis, Investigation, Resources, Visualization, Project Administration, Writing–original draft, Writing–review & editing.

**Saul Diaz-Infante:** Conceptualization, Methodology, Software, Validation, Formal analysis, Investigation, Data curation, Visualization, Supervision, Writing–original draft, Writing–review & editing.

**David Baca-Carrasco:** Conceptualization, Methodology, Formal analysis, Writing–original draft, Writing–review & editing.

**Daniel Olmos Liceaga:** Conceptualization, Methodology, Formal analysis, Writing–original draft, Writing–review & editing.

## Acknowledgments

The authors acknowledge support from grant DGAPA-PAPIIT IV100220 and the Laboratorio Nacional de Visualización Científica UNAM. MAAZ acknowledges support from PRODEP Programme (No. 511-6/2019-8291). DBC acknowledges support from PRODEP Programme (No. 511-6/2019-8022). We thank Dr. Jorge X. Velasco-Hernandez for useful comments and discussions of this work.

## A. Parameter estimation

Mathematical models for COVID-19 have shown that the parameters’ values are not necessarily the same in each country. We use COVID-19 data from Mexico City plus Mexico state to follow the epidemic curve’s initial growth in this work. Consequently, we estimate some parameter values of system in Equation (1) [30]. To obtain the baseline parameter values, we consider two-stages: i) before and ii) after mitigation measures were implemented. For both stages, we use model in Equation (1) with no vaccination dynamics (*λ*_*V*_ = 0 and *V* (0) = 0), and STAN R-package. This package is used for statistical inference by the Bayesian approach. For the code implementation of our system, we follow ideas of [46], and it is made freely available at [39]. For this section, our estimations are focused on three parameters: *β*_*A*_, *β*_*S*_ and *p*. Other parameter values are given in Table A.7.

**Table A.7:** Fixed parameters values of system in Equation (1).

| Parameter | Value | References |
| --- | --- | --- |
| $\delta_E^{-1}$ | 5.1 days | [47] |
| $\alpha_S^{-1}$ | 5.97 days | [10] |
| $\alpha_A^{-1}$ | 10.81 days | [10] |
| $\delta_R^{-1}$ | 365 days | |
| $\mu^{-1}$ | 70 years | |

For the first stage, the following system is considered:

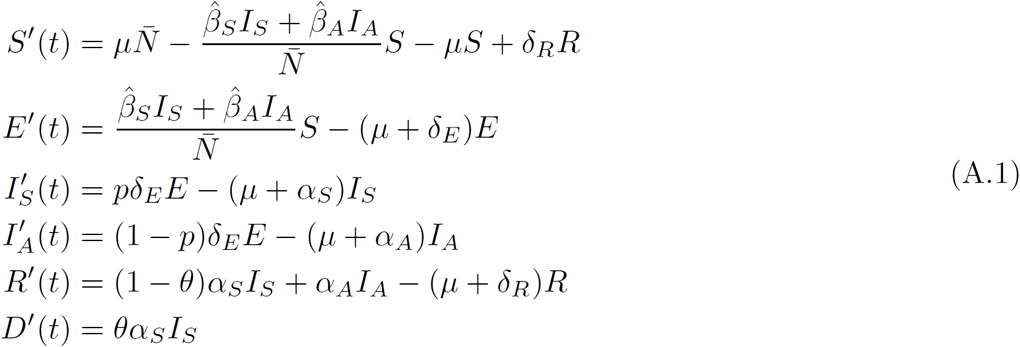

where 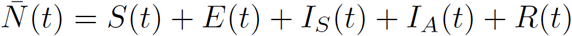 and 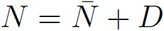. Here, we consider COVID-19 data from the first day of symptoms onset reported (February 19) until March 23, 2020. We also assume that *θ* = 0 because the first reported death was on March 18, and there were three reported deaths until March 23. The initial values of recovered and dead people are set to zero. Symptomatic class initial value was fixed in one individual, while *E*(0) and *I*_*A*_(0) were estimated. Thus, *S*(0) = *N* − (*E*(0)+*I*_*A*_(0)+ 1), where *N* = 26446435 [48]. For the STAN implementation, we employ a negative-binomial model as the likelihood function with the mean parameter given by incidence solution per day. In addition to the above, we assign prior probability distributions to each parameter and the exposed and asymptomatic classes’ initial conditions. Thus, we propose that 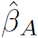 and 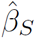 follow a normal distribution with parameters *µ* = 1 and *σ*^2^ = 0.13. Then, *p* follows a uniform distribution in (0, 0.25), and *E*(0) and *I*_*A*_(0) also follow a uniform distribution in (2, 20) and (2, 10), respectively. When employing our STAN implementation, we run 5 chains with 100,500 iterations each, discard the first 500, and use 10,000 samples to generate estimates of parameters 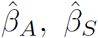, and *p*. Table A.8 shows the confidence interval for each parameter and median posterior estimated.

**Table A.8:** Confidence interval and median posterior estimated for some parameters of system in Equation (A.1) and basic reproductive number (*R*_0_).

| Parameter | 95% Confidence Interval | Quantile 50 |
| --- | --- | --- |
| $\hat{\beta}_S$ | [0.672, 1.1886] | 0.9322 |
| $\hat{\beta}_A$ | [0.501, 0.7851] | 0.6435 |
| $p$ | [0.061, 0.2206] | 0.1227 |
| $R_0$ | [4.159, 5.1991] | 4.6082 |

For the second stage, we took a complete month starting the day when mitigation measures were implemented, that is, from March 23 to April 23, 2020. Now, we consider parameter *ξ* to model the implementation of non-pharmaceutical measures. Thus, system in Equation (A.1) becomes:

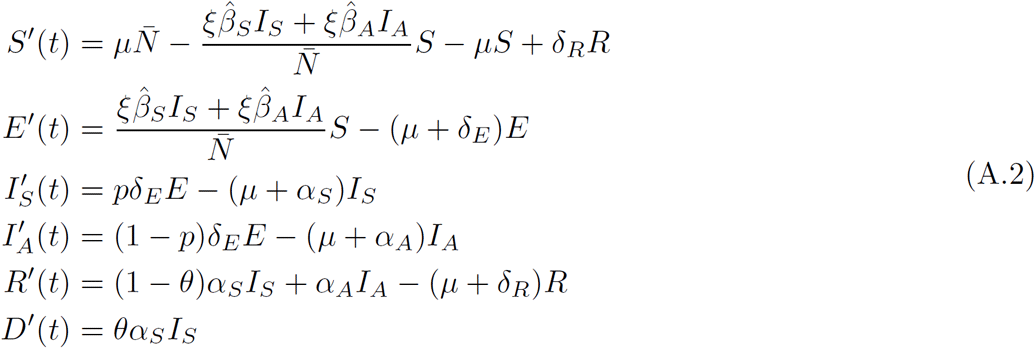

where 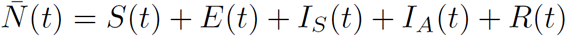 and 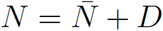. At this stage, we consider that *θ* = 0.11. Here, our objective is to estimate the value of parameter *ξ*. To do this, we use the median posterior of all the estimated parameters from the first stage (see Table A.8). Other parameter values are given in Table A.7. Almost all initial conditions were obtained when solving system in Equation (A.1) with the 10,000 samples (obtained in first stage), after which each solution at the final time (March 23) is saved. We use the median of the saved values. Thus, for system in Equation (A.2), *E*(0) = 6587.585, *I*_*S*_(0) = 553.7035, *I*_*A*_(0) = 3149.924, and *R*(0) = 3001.547. For the initial value of variable *D*, we consider reported COVID-19 data, then *D*(0) = 3. Therefore *S*(0) = *N* − (*E*(0) + *I*_*S*_(0) + *I*_*A*_(0) + *R*(0) + *D*(0)), with *N* = 26446435 [48]. Similar to the first stage, we consider a negative-binomial model as the likelihood function with the mean parameter given by incidence solution per day, while that we postulate a uniform distribution in (0.25, 0.75) as a prior probability distribution for the parameter *ξ*. For the second stage, we run 5 chains with 100,500 iterations each, discard the first 500, and use 10,000 samples to generate estimates of parameters *ξ*.

**Table A.9:**
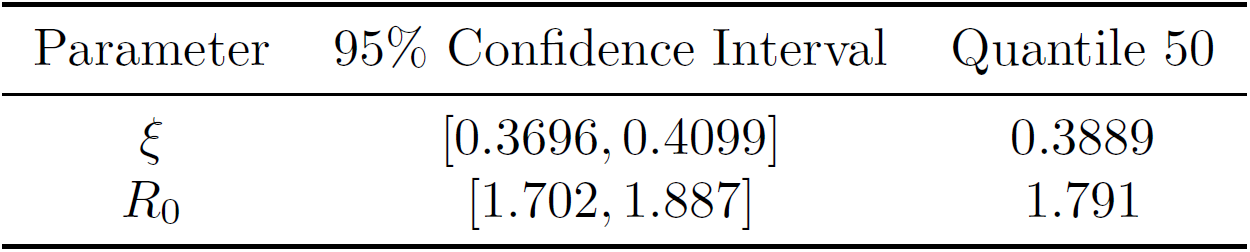
Confidence interval and median posterior estimated for parameter *ξ* of system in Equation (A.2) and basic reproductive number (*R*_0_).

| Parameter | 95% Confidence Interval | Quantile 50 |
| --- | --- | --- |
| $\xi$ | [0.3696, 0.4099] | 0.3889 |
| $R_0$ | [1.702, 1.887] | 1.791 |

Finally, it is important to mention that our results were implemented considering that the effective transmission contact rates (*β*_•_) were equal to 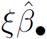. This last means that our scenarios consider the first reduction in the effective transmission contact rates by NIPs. Using values in Tables A.8 and A.9, we build confidence intervals for 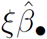. These results are shown in Table A.10.

**Table A.10:** Confidence interval for parameters *β*_*•*_.

| Parameter | 95% Confidence Interval |
| --- | --- |
| $\beta_S = \xi\hat{\beta}_S$ | [0.2483712, 0.48720714] |
| $\beta_A = \xi\hat{\beta}_A$ | [0.1851696, 0.32181249] |

## B. Positivity and Invariance of the Interest Region

### Lemma 1.

*The set* 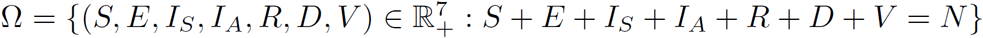 *is a positively invariant set for the system in* *Equation* (1).

*Proof*. Let 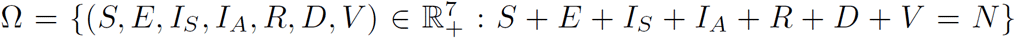. First, note that for this model we have a closed population, which allows the solutions to be bounded superiorly by the total population.

On the other hand, to show the positivity of the solutions with initial conditions

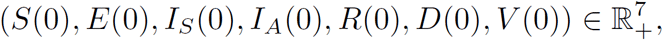

we look at the direction of the vector field on the hypercube faces in the direction of each variable in the system. For example, consider a point on the hypercube face where the variable *S* = 0 and look at the behavior of the vector field in the direction of the same variable *S*, to see if the solutions cross the face of the hypercube where we are taking the initial condition. So, notice that if *S* = 0, *S*^*′*^(*t*) *>* 0, so the solution points into the hypercube. Similarly, consider an initial condition of the form (*S*, 0, *I*_*S*_, *I*_*A*_, *R, D, V*) and note that *E*^*′*^ (*t*) *>* 0 for all *t >* 0, which implies that the solutions of the system with initial conditions of the form (*S*, 0, *I*_*S*_, *I*_*A*_, *R, D, V*) point towards the interior of the hypercube. Similarly, positivity can be tested for the rest of the variables. With this information, we have the following result.□

Now, continuing with the analysis of our model, it is easy to prove that the disease-free equilibrium is given by the point at *X*_0_ *∈* Ω of the form

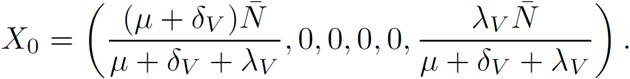

On the other hand, following ideas of [25], the next generation matrix for this model, evaluated in the disease equilibrium point, is given by

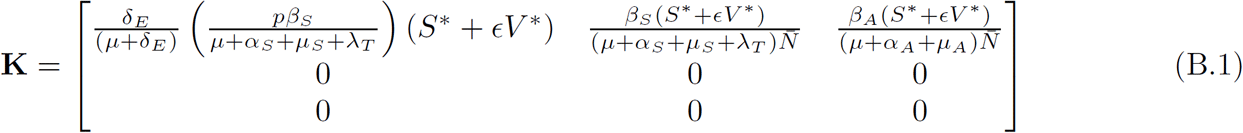

where 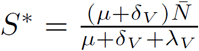 and 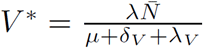. Then, the spectral radius of **K** is

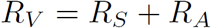

with

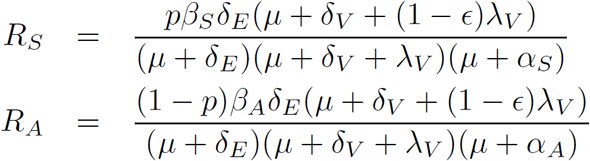

